# Association between vaccination rates and COVID-19 health outcomes in the United States: a population-level statistical analysis

**DOI:** 10.1101/2023.06.14.23291388

**Authors:** Hongru Du, Samee Saiyed, Lauren M. Gardner

**Affiliations:** Center for Systems Science and Engineering, Johns Hopkins University, Baltimore, MD 21218, USA; Department of Civil and Systems Engineering, Johns Hopkins University, Baltimore, MD 21218, USA; Department of Epidemiology, Johns Hopkins Bloomberg School of Public Health, Baltimore, MD, 21205, USA

**Keywords:** COVID-19, vaccination rates, population-level, United States, statistical analysis

## Abstract

Population-level vaccine efficacy is a critical component of understanding COVID-19 risk, informing public health policy, and mitigating disease impacts. Unlike individual-level clinical trials, population-level analysis characterizes how well vaccines worked in the face of real-world challenges like emerging variants, differing mobility patterns, and policy changes. In this study, we analyze the association between time-dependent vaccination rates and COVID-19 health outcomes for 48 U.S. states. We primarily focus on case-hospitalization risk (CHR) as the outcome of interest, using it as a population-level proxy for disease burden on healthcare systems. Performing the analysis using Generalized Additive Models (GAMs) allowed us to incorporate real-world nonlinearities and control for critical dynamic (time-changing) and static (temporally constant) factors. Dynamic factors include testing rates, activity-related engagement levels in the population, underlying population immunity, and policy. Static factors incorporate comorbidities, social vulnerability, race, and state healthcare expenditures. We used SARS-CoV-2 genomic surveillance data to model the different COVID-19 variant-driven waves separately, and evaluate if there is a changing role of the potential drivers of health outcomes across waves. Our study revealed a strong and statistically significant negative association between vaccine uptake and COVID-19 CHR across each variant wave, with boosters providing additional protection during the Omicron wave. Higher underlying population immunity is shown to be associated with reduced COVID-19 CHR. Additionally, more stringent government policies are generally associated with decreased CHR. However, the impact of activity-related engagement levels on COVID-19 health outcomes varied across different waves. Regarding static variables, the social vulnerability index consistently exhibits positive associations with CHR, while Medicaid spending per person consistently shows a negative association. However, the impacts of other static factors vary in magnitude and significance across different waves. This study concludes that despite the emergence of new variants, vaccines remain highly correlated with reduced COVID-19 harm. Therefore, given the ongoing threat posed by COVID-19, vaccines remain a critical line of defense for protecting the public and reducing the burden on healthcare systems.

## 1. Background

By March 1^st^, 2023, the COVID-19 pandemic caused over 102 million reported cases and 1.1 million deaths in the United States. Vaccine development and distribution have been at the forefront of efforts to combat the impact of the disease. Three vaccines are currently available in the U.S., developed by Pfizer-BioNTech, Moderna, and Johnson & Johnson. Initial randomized clinical trials demonstrated the safety and efficacy of these vaccines, with vaccine efficacies against severe disease (hospitalization and death) ranging from 73.1% to 96.7% [1–3]. The clinical trials were designed to estimate the direct effect of vaccines against severe disease at the individual level [4]. However, as vaccines roll out to a broader population, uncertainties such as the emergence of new variants, variable immune responses, the quality of cold-chain storage, and other confounding factors can impact a vaccine’s efficacy [5]. Hence, evaluating real-world vaccine protection against COVID-19 health outcomes poses a challenge.

Several published studies have attempted to quantify the real-world impact of the COVID-19 vaccines on health outcomes. For example, a study in Qatar assessed the vaccines’ effectiveness against severe, critical, or fatal Omicron infections using test-negative case-control analysis, and found previous infections and vaccination are effective against symptomatic Omicron infections [6]. An observational study conducted in Israel using national surveillance data showed that the two doses of the Pfizer-BioNTech mRNA vaccines are 97.2% effective in preventing COVID-19-related hospitalizations [7]. A Danish study estimated vaccine effectiveness against COVID-19 hospitalization using a cohort study design, and found that two doses of the vaccine provide high protection against hospitalization for the Alpha and Delta variant, and even higher protection against hospitalization for the Omicron variant [8]. A similar cohort study was applied in Singapore and the United Kingdom to determine whether booster shots reduce the severity of COVID-19 infections during the Omicron wave, and found consistent results that the risk of severe COVID-19 outcomes reduced after receiving booster mRNA vaccines [9,10].

Most existing literature on the population-level effects of COVID-19 vaccination is based on individual-level data and observational studies. Specifically, these studies relied upon detailed individual-level data to assess the direct effectiveness of vaccination by comparing health outcomes between vaccinated and unvaccinated individuals exposed to the same environment. However, these studies may be subject to confounding by unmeasured factors and inconsistent quality of individual-level data. Further, in the U.S., such high-resolution data is unavailable at the population-level, so alternative strategies must be engaged to evaluate the impact of vaccine at a regional level.

One such approach is to rely on compartmental and agent-based models to simulate transmission and disease outcomes both in the presence and absence of vaccines implementation for the same population. Watson et al applied this method to estimate the impact of varying vaccine uptake rates on mortality across multiple countries and found that vaccines prevented 14·4 million COVID-19 deaths in 2021 [11]. However, this approach is subject to many assumptions and is limited in its ability to estimate accurate effectiveness. Alternatively, statistical methods such as time series and regression analysis can be implemented to evaluate the association between vaccination coverage and healthcare outcomes across different locations. One study using this strategy evaluated the association between vaccination coverage and the COVID-19 cases growth rate for all 50 U.S. states in the U.S. using a structural nested mean model and found a 1% increase in vaccination coverage was associated with a 1.02% reduction in case growth rate [12]. However, the scope of this study is limited to cases between March and May 2021. Another study utilized linear regression to analyze vaccine coverage and natural immunity in relation to mortality during the Delta and Omicron waves. It found that vaccine coverage reduced COVID-19 mortality, but seroprevalence and prior infection rates were not associated with mortality [13].

However, this method has limitations in capturing dynamic changes and non-linear relationships between variables. A different study by Bollyky et al [14] applied regression analysis to determine how vaccination coverage amongst other factors (e.g., presence of comorbidities, political partisanship, race, and ethnicity) impacted health outcomes (standardized infections and deaths) in the U.S. at the state level, and determined that higher vaccination rates were associated with lower death rates. The scope of this study varies from ours in its focus on the association between static variables and COVID-19 health outcomes for a fixed time window between January 1^st^, 2020, and July 31^st^, 2022, while our study expands the analysis by incorporating novel dynamic variables to capture behavioral changes over time, and explicitly evaluating the different variants independently. A recent study evaluated the time-varying relationship between vaccination, mobility, and COVID-19 health outcomes before and after the Omicron waves [15]. They found the significance of the vaccine’s impact in reducing case rates diminished during the Omicron surge, while its efficacy in lowering case-fatality rates remained substantial throughout the pandemic.

Our study contributes to the existing literature by prioritizing case-hospitalization risk as the outcome variable, breaking aggregated mobility into activity-related engagement levels, modeling previous infections as a dynamic variable, including an interaction between the completed primary series and booster rate for the Omicron wave, and considering the critical static factors such as comorbidities, social vulnerability, race, and state healthcare expenditures. Despite numerous studies assessing the effectiveness of vaccines, most have not accounted for the relative impact of vaccines across different populations and variant waves, while considering dynamic potential confounding factors. Therefore, a more comprehensive understanding of vaccines’ impact across diverse populations and COVID-19 waves is crucial in developing informed public health policies that can effectively mitigate the spread of the virus and ensure equitable distribution of healthcare resources.

## 2. Methods

### 2.1 Study design

The primary objective of this study is to analyze the association between COVID-19 vaccination rates and COVID-19 case-hospitalization risk (CHR) in the U.S. while controlling for potential confounding effects. Time-dependent COVID-19 CHR is chosen as the modeled response variable to gain insights into the factors influencing COVID-19 harm; CHR serves as both a proxy for disease severity at an individual level, and captures the burden on the healthcare system at a population level. We use Generalized Additive Models (GAMs) to perform the analysis because of their ability to capture nonlinear dynamics. Data used include novel dynamic covariates that may potentially contribute to COVID-19 CHR, such as naturally derived immunity from prior COVID-19 infection, local healthcare infrastructure, activity-related engagement levels in the population, and government policies, alongside various static variables that have been identified to be significant in previous studies [14,16,17] such as comorbidities, social vulnerability index (SVI), race, and state healthcare expenditures. By controlling for these factors, we aim to provide a more comprehensive understanding of the association between vaccination rate and COVID-19 CHR at the population level. To further elucidate the role of potential driving factors, we also model reported case incidence rates (CIR) as a separate response variable and compared the factors associated with COVID-19 transmission versus those associated with COVID-19 CHR. Our framework explicitly captures the spatial variation in the modeled relative associations through a variable transformation procedure (discussed in detail in the methods section). The study was conducted for 48 states in the U.S. for the period between April 19^th^, 2021, the date at which the vaccines were approved for all adults in the U.S., to March 1^st^, 2022. This period covers the pre-Delta (characterized by the predominance of the Alpha variant and other variants), Delta, and Omicron waves of COVID-19, which are each evaluated independently. To distinguish between COVID-19 variant-driven waves, we utilized SARS-CoV-2 genomic surveillance data and identified the dominant variant for each state and point in time, to determine time windows so the distinct variant-driven waves can be modeled independently. For the Omicron wave, we also considered the added benefit of booster doses on COVID-19 health outcomes. Specifically, we evaluated the interaction between the completed primary series and booster rate on reducing COVID-19 CHR. Results from this analysis help improve our understanding of the real-world relative impact of the available COVID-19 vaccines against COVID-19 CHR at the population-level over time, and can help inform future public health policies to reduce harm.

### 2.2 Data sources and collection

We collected state-level time-series data and static variables from publicly available databases. All time-series data were aggregated to the weekly level. A summary of the variables and their respective sources are listed in Table 1, and detailed explanations of each variable are provided in Appendix section 1.2. A 3-week moving average was applied to all time-series variables to mitigate the effects of potential noise and reporting issues, with the exception of the government policy index.

**Table 1.** Summary of dynamic variables in the model.

| Variable Name | Variable Description | Source |
| --- | --- | --- |
| **Output variables** |  |  |
| Case-hospitalization rate (CHR) | Weekly new admissions of patient with confirmed COVID-19 normalized by reported cases for each state. | [18], [19] |
| Reported case-incidence rate (CIR) | Weekly number of confirmed cases normalized by state population. | [18] |
| **Dynamic input variables** |  |  |
| Partial vaccination rate | Percentage of the total population that received at least one dose of COVID-19 vaccine approved or authorized for use in the United States. | [20] |
| Completed primary series rate | Percentage of the population that received the second dose in a two-dose COVID-19 vaccines primary series or one dose of a single-dose COVID-19 vaccine primary series approved or authorized for use in the United States. | [20] |
| Booster vaccination rate | Percentage of the total population that received an updated (bivalent) booster dose. | [21] |
| Weekly testing rate | Total number of weekly tests conducted for each state normalized by population. | [22] |
| Gym visits | Number of weekly visits to gyms per person. | [23,24] |
| University visits | Number of weekly visits to universities per person. | [23,24] |
| Physician visits | Number of weekly visits to physicians per person. | [23,24] |
| Government policy index | Quantitative measure of government policies implemented in response to the COVID-19 pandemic across various domains including health, social, and economic policies. | [25] |
| Previous infections | Infections reported within a time window preceding the modeled output, e.g., sum from 4 to 16 weeks ahead of the output variables. | [18] |

**Table 2.** Summary of static variables in the model.

| Static input variables | Description | Mean | St.d. | Min | Max | Source |
| --- | --- | --- | --- | --- | --- | --- |
| Black proportion | The proportion of the population identified as Black. | 0.112 | 0.019 | 0.006 | 0.378 | [26] |
| Social vulnerability index (SVI) | The Social Vulnerability Index utilizes data from the U.S. Census to assess the relative level of social vulnerability in each census tract. By analyzing 14 social factors, the SVI categorizes tracts into four closely interrelated themes and then aggregates them as a single indicator of social vulnerability. | 0.468 | 0.152 | 0.137 | 0.771 | [27] |
| Adults at high risk | The proportion of the population over 18 years old is at high risk of serious illness if infected with Coronavirus. | 0.383 | 0.036 | 0.300 | 0.493 | [28] |
| Medicaid spending | Total Medicaid spending in thousands of dollars for each state normalized by the population. | 1.807 | 0.550 | 0.860 | 3.099 | [29] |

### 2.3 Dynamic variable transformation

To ensure the precise estimation of each dynamic variable’s impact, a variable transformation mechanism must be used to account for the effects of time trends in the data. For example, the completed primary series rate is always increasing with time for all locations modeled, hence it can be difficult to distinguish how much of the observed associations between vaccination rate and COVID-19 health outcomes are due to the variable interaction or the passage of time. Moreover, the main focus of this study lies in modeling spatial differences and considering location-specific variations that influence the observed associations. Consequently, we applied the following transformation to all dynamic variables to remove the time trend and redefine the relative variable (RV^t^):

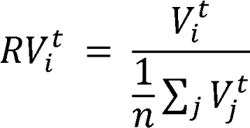

Where RV_i_^t^ represents the transformed variable for state *i* at week t, V_i_^t^ represents the original variable for state i at week t without the transformation, 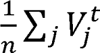 represents the mean of the original variable at week t, over all locations being modeled *n*, e.g., the national mean across the U.S. A RV_i_^t^ larger than one indicates that state i has a higher variable value compared to the national mean at week *t*, while RV_i_^t^ lower than one indicates that state i has a lower variable value compared to the national mean, at week *t*. After normalization, the final set of time-dependent variables included in the analysis are: Relative case-hospitalization rates (RCHR_i_^t^), relative reported case-incidence rate (RC/R_i_^t^), relative completed primary series rate (RCPSR_i_^t^), relative booster rate (RBR_i_^t^), relative weekly testing rate (RWTR_i_^t^), relative gym visits (RGV_i_^t^), relative physician visits (RPV_i_^t^), relative university visits (RUV_i_^t^), relative previous infection (RP_i_^t^), and relative government policy (RGP_i_^t^). These newly transformed variables enable an explicit evaluation of the relative association between each of them and the COVID-19 health outcome of interest within a single multi-state model. Moreover, this variable transformation procedure facilitates assessing individual state’s performance relative to national dynamics. It emphasizes evaluating the expected outcomes when a state’s performance diverges from the national average.

The dynamic variables, with and without variable transformation, are visually depicted in Appendix figure S2. Among all the variables, the rankings of RCPSR^t^ remain relatively stable across time, as seen in Appendix figure S2b2. This stability indicates a more consistent spatial-temporal pattern of variation among vaccination rates across states. On the other hand, all other dynamic variables exhibit more noticeable spatial ranking changes over time. The changing spatial-temporal rankings of other dynamic variables highlight the importance of considering spatial differences through time and evaluating their influence on COVID-19 health outcomes.

### 2.4 Statistical analysis

The generalized additive model (GAM) was selected as the statistical model for this analysis because of its ability to capture complex and nonlinear relationships between the set of covariates and the outcome variables of interest in each state. We independently model each variant-driven wave during the study period to allow for different driving factors for different variants. To define the variant waves, we clustered each state-week pair based on the dominant circulating variant based on SARS-CoV-2 genomic surveillance data downloaded from GISAID [30]. The three waves are classified as: 1) Pre-Delta Wave, 2) Delta Wave, 3) Omicron Wave, and each state is labeled with its most dominant variant each week to define the windows. Details of this classification are described in Appendix section 1.1, and the assignment of state-week pairs is shown in Appendix figure S1.

The primary set of models treat weekly state-level RCHR as the response variable, with separate models generated independently for each variant wave, namely Pre-Delta-RCHR, Delta-RCHR, and Omicron-RCHR. These three models have the form:

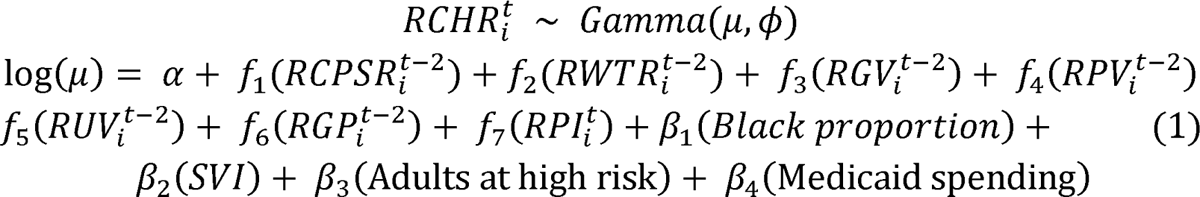

Where α represents the intercept, β_i_ represent the parametric coefficients of each static variable, and *f_i_* are spline smooth functions of the relative dynamic variables. Additionally, a model is constructed for the Omicron wave, incorporating an interaction between completed primary series and booster rate (Omicron-Booster-RCHR). The Omicron-Booster-RCHR has the form:

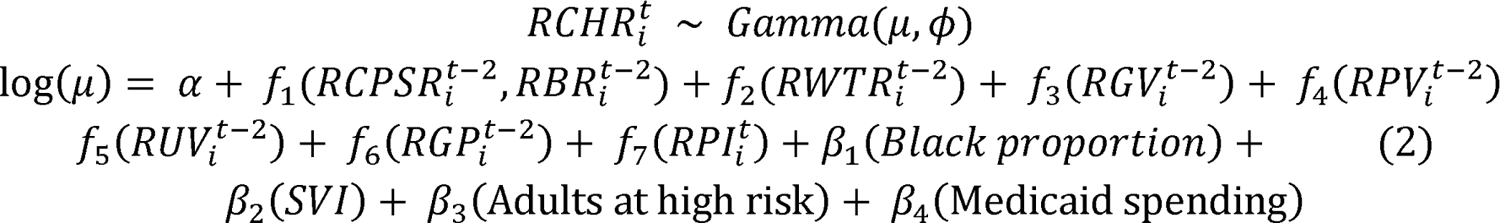

Where *f_1_* represent a smooth interaction function between RCPSR_i_^t-2^ and RBR_i_^t-2^. For all the mentioned models above, the weekly state-level RCHR is assumed to follow a Gamma distribution with a log link. This choice of the Gamma family accounts for the positively skewed distribution of the outcome variable. We use thin plate regression splines as the smoothing basis for all *f*_i_ and set the basis dimension to three to maximize the interpretability of the models. The basis dimension refers to the maximum possible complexity of each smooth term; a large basis dimension could overfit the data and result in highly non-linear relationships between input and outcome variables.

To consider the sequential process of infection leading to hospitalization we introduce a time lag between each of the input variables relative to the outcome variable, which is denoted by the superscript. The timeline of this model is introduced as follows: the modeled relative case-hospitalizations rate (RCHR_i_^t^), occur at time *t*. Infections resulting in hospitalization, are assumed to occur at time *t-2*, to account for a one week incubation period [31], and one additional week between symptoms onset and hospitalization [32]. Note, this timeline aligns with the definition of the CHR variable, which is normalized by the number of reported infections one week prior, which assumes a one week delay between when infection occurred and when it is reported. To accurately reflect the conditions presented at the time of infection, each of the variables related to vaccination (RCPSR_i_^t-2^), activity-related engagememnt levels (RGV_i_^t-2^, RPV_i_^t-2^, RUV_i_^t-2^), policy (RGP_i_^t-2^), and testing (RWTR_i_^t-2^) are also lagged by two weeks relative to the case-hospitalization risk. Lastly, the past infections variable, defined as stated above to capture the role of recently acquired immunity from infection in protecting from severe disease upon reinfection, is equal to the total infection rate in the population summed over the prior 4 to 16 weeks. This time window is explicit in the definition of RP/^t^ (see Appendix section 1.2).

A secondary set of analogous models treats RCIR as the response variable, namely the Pre-Delta-RCIR, the Delta-RCIR, the Omicron-RCIR, and the Omicron-Booster-RCIR. The first three models adopt the same form as equation (1), while the Omicron-Booster-RCIR follows the same form as equation (2). To account for the sequential process leading to infections, all lag between dynamic covariates, and RCIR have been reduced by one week. This results in eight models, with four models fit to RCHR, and four models fit to RCIR. The exact formulation of models with RCIR as outcome variable are documented in Appendix section 2.6.

The selection of covariates for each model relies on correlation-based feature selection, taking into account both Pearson’s correlation between variables and the concurvity measures derived from GAMs. Details regarding feature selection can be found in Appendix sections 2.1 and 2.2. The impact of each dynamic variable is quantified by computing the Accumulated Local effect (ALE) of each smooth term on outcome variables. The local effect refers to the change in model output when a particular input feature is changed while keeping all other features constant. The ALE method aggregates the local effects of each input feature across its entire range. By accumulating these local effects, we gain insight into how changes in each input variable influence the outcome variable across its entire range. Data processing, visualization, and analysis were carried out using R 4·0 and Python 3·8.

## 3. Results

### 3.1 GAMs analysis for the relative case-hospitalization rate (RCHR) as the outcome

In our analysis, we evaluated goodness-of-fit based on several metrics. For models with relative case-hospitalization risk (RCHR) as the outcome variable, the deviance explained ranges between 46.8% and 72.3% (Figure 1l) for each variant wave. Moreover, we assessed the correlation between observed RCHR and predicted RCHR, which exhibited strong positive correlations ranging from 0.67 to 0.81 (Appendix section 3.1). These findings provide compelling evidence of the models’ effectiveness in capturing and predicting the case-hospitalization rate.

**Figure 1:**
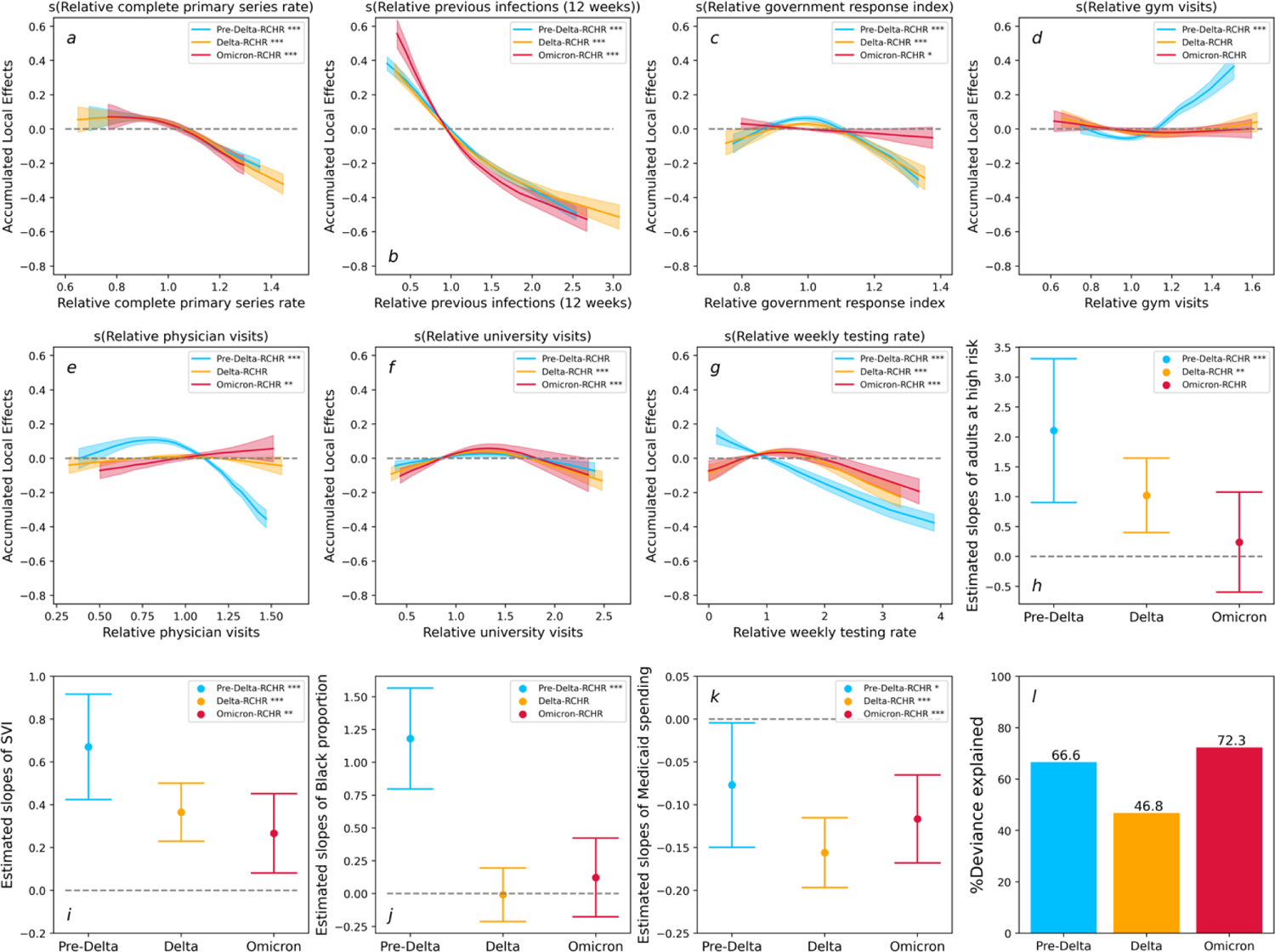
Results for the Pre-Delta-RCHR (Blue), the Delta-RCHR (Orange), and the Omicron-RCHR (Red). a-g: Accumulated local effects (ALE) of dynamic variables. Shaded areas in each plot indicate 95% confidence intervals. h-k: Estimated slopes for each static variables, the upper and lower band indicate 95% confidence intervals. l: Deviance explained for each model. ‘***’: variable significant at p<0.001. ‘**’: variable significant at p<0.01. ‘*’: variable significant at p<0.05. ‘’: variable significant at p>0.05.

The relative completed primary series rate, and relative previous infections consistently displayed strong negative associations with RCHR across different waves (Figures 1a and 1b). Of particular note is that relative previous infections consistently ranked the highest in terms of ALE across the different waves. Figure 1c reveals the impact of the relative government response index gradually flattening out from the pre-Delta to the Omicron wave. Regarding activity-related engagement levels, their effects on RCHR appear inconsistent across different waves, as exemplified by the relative physician visits, which slightly changed from negative to positive effects as the analysis progressed from the pre-Delta to the Omicron wave (Figure 1e). Lastly, the relative weekly testing rate served as a control variable to address the state-level differences in testing rates. The result revealed a negative correlation between the relative weekly testing rate and RCHR. Nevertheless, it is noteworthy that this association exhibited a decrease from the pre-Delta wave to the Omicron wave, as illustrated in Figure 1g.

Regarding the static variables, adults at high risk exhibited a declining positive association with RCHR. Additionally, states with higher Social Vulnerability Index (SVI) consistently showed higher RCHR. Among racial groups, the proportion of Black positively associated with RCHR during the Pre-Delta wave but did not exhibit a significant impact since the Delta wave. With healthcare systems variables, Medicaid spending per person consistently showed a negative association with RCHR.

With the exception of the completed primary series rate, the effects of all other variable modeled in the Omicron-Booster-RCHR remained consistent with the results for the Omicron-RCHR shown in Figure 1. Figures 2a show the interaction between two vaccine-related variable in a two-dimensional variable space. The solid black lines represent the contour lines. The contour lines correspond to points that have an equivalent impact on the hospitalization rate, with the values marked on each line indicating the actual interaction effect of these points on the RCHR. Figure 2a reveals that the RCHR decreases along the direction of increasing the relative booster rate and the relative completed primary series rate.

**Figure 2:**
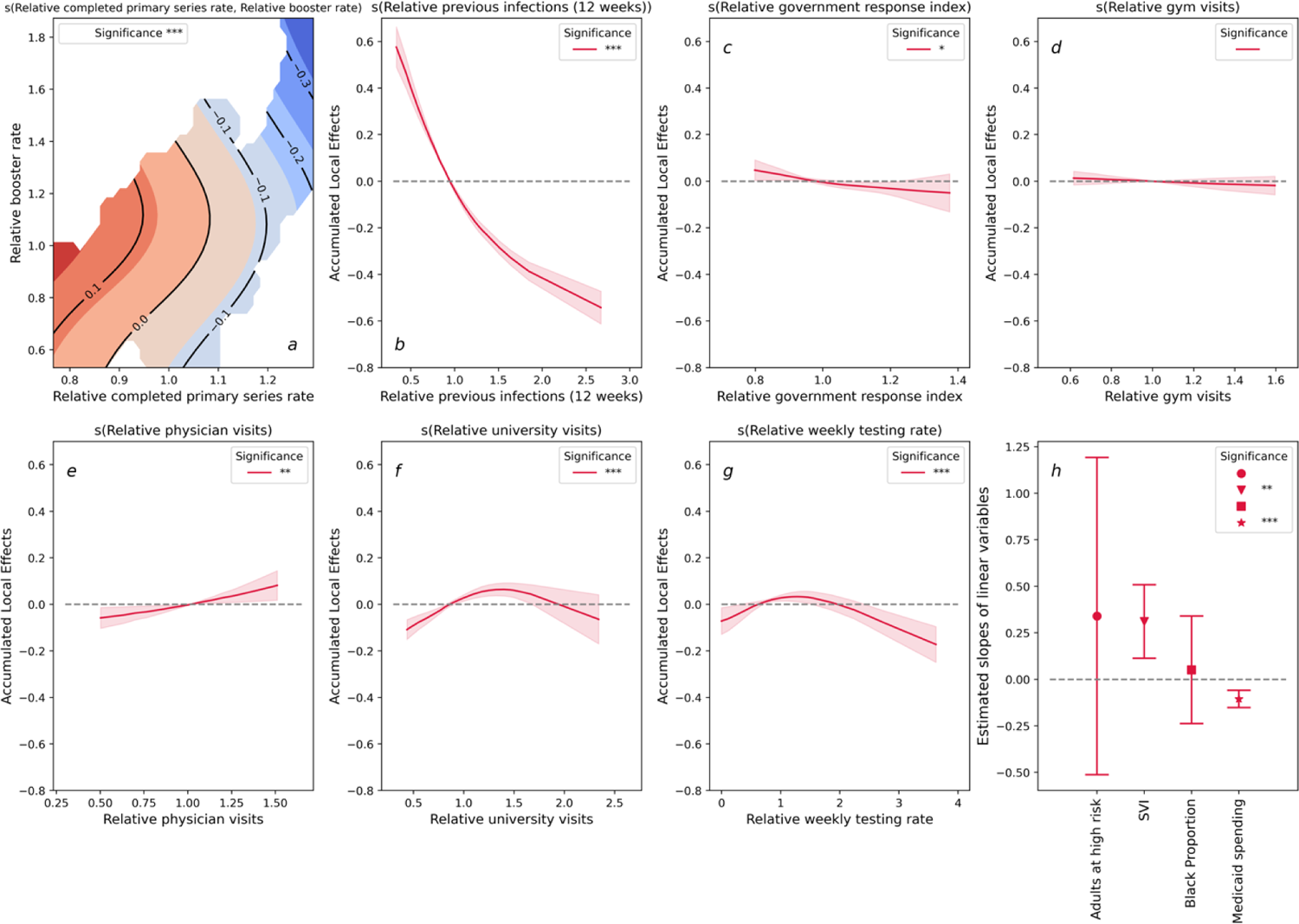
Results of Omicron-Booster-RCHR for just the Omicron wave with the additional inclusion of an interaction effect between the relative completed primary series rate and the relative booster rate. a: Two-dimensional contour plot for the interaction between relative completed primary series rate and relative booster rate. The deeper red indicates a more positive effect on the RCHR, and the deeper blue indicates a more negative effect to the RCHR. b-g: Accumulated local effects (ALE) of dynamic variables. Shaded areas in each plot indicate 95% confidence intervals. h: Estimated slopes for each static variables, the upper and lower band indicate 95% condifence intervals. ‘***’: variable significant at p<0.001. ‘**’: variable significant at p<0.01. ‘*’: variable significant at p<0.05. ‘’: variable significant at p>0.05.

### 3.2 GAMs analysis for the relative reported case-incidence rate (RCIR) as the outcome

The GAMs using relative reported case-incidence rate (RCIR) as the outcome variable consistently demonstrate lower performance than those GAMs with RCHR as the outcome variable. Specifically, all GAMs for RCIR have deviance explained values below 40%, and correlations between observed RCIR and predicted RCIR range from 0.43 to 0.61 (Appendix section 3.2). The observed performance pattern indicates a more intricate and dynamic relationship concerning COVID-19 transmission, particularly evident during the Omicron wave.

Figure 3a illustrates a strong negative association between the relative completed primary serie rate and RCIR during the Pre-Delta and Delta waves. However, this association vanished during the Omicron wave, coinciding with a decline in model performance (Figure 3l). The ALE plot of the relative previous infection rate (Figure 2b) revealed an insignificant association between previous infection and RCIR during the pre-Delta and Delta waves but a significant negative association during the Omicron wave. Additionally, when the relative government policy index is greater than one, the ALE plots demonstrate a negative trend; however, the magnitude of this effect is relatively smaller compared to other dynamic variables examined in the analysis. Similar to GAMs for RCHR, the activity-related engagement levels exhibited inconsistent patterns across different waves. Notably, the ALE of relative university visits reverses direction from negative to positive between the pre-Delta wave and the later two waves.

**Figure 3:**
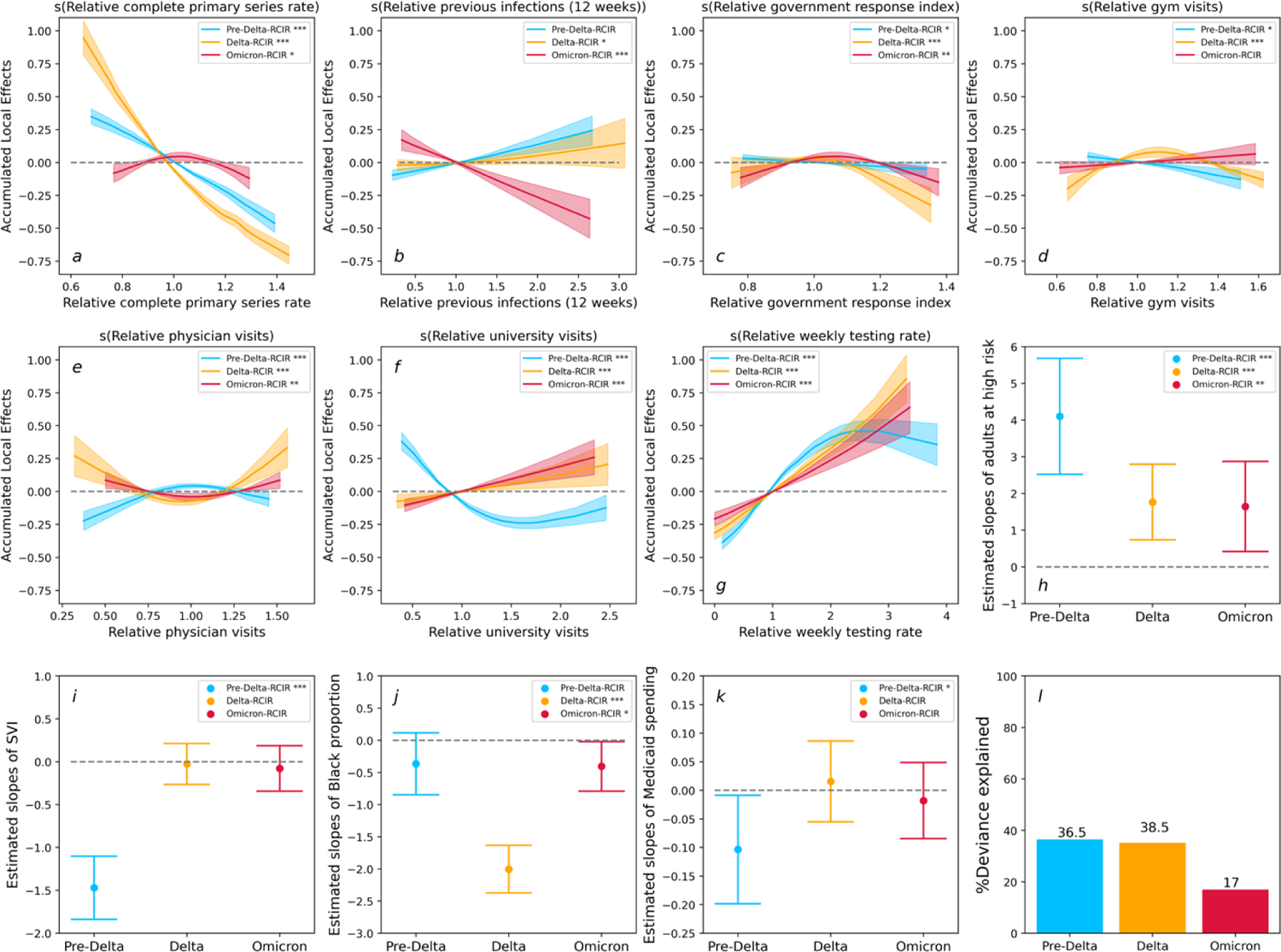
Results for the Pre-Delta-RCIR (Blue), the Delta-RCIR (Orange), and the Omicron-RCIR (Red). a-g: Accumulated local effects (ALE) of dynamic variables. Shaded areas in each plot indicate 95% confidence intervals. h-k: Estimated slopes for each static variables, the upper and lower band indicate 95% condifence intervals. l: Deviance explained for each model. ‘***’: variable significant at p<0.001. ‘**’: variable significant at p<0.01. ‘*’: variable significant at p<0.05. ‘’: variable significant at p>0.05.

For the static variables, adults at high risk were consistently positively associated with RCIR across different waves. However, the other static variables, including racial groups, SVI, and healthcare expenditures, do not show a consistent or significant impact across different waves.

Figure 4 illustrates the results of the Omicron-Booster-RCIR for just the Omicron wave with the additional inclusion of an interaction effect between the completed primary series rate and the relative booster rate. This interaction effect is presented as a dimension contour map in figure 4a.

**Figure 4:**
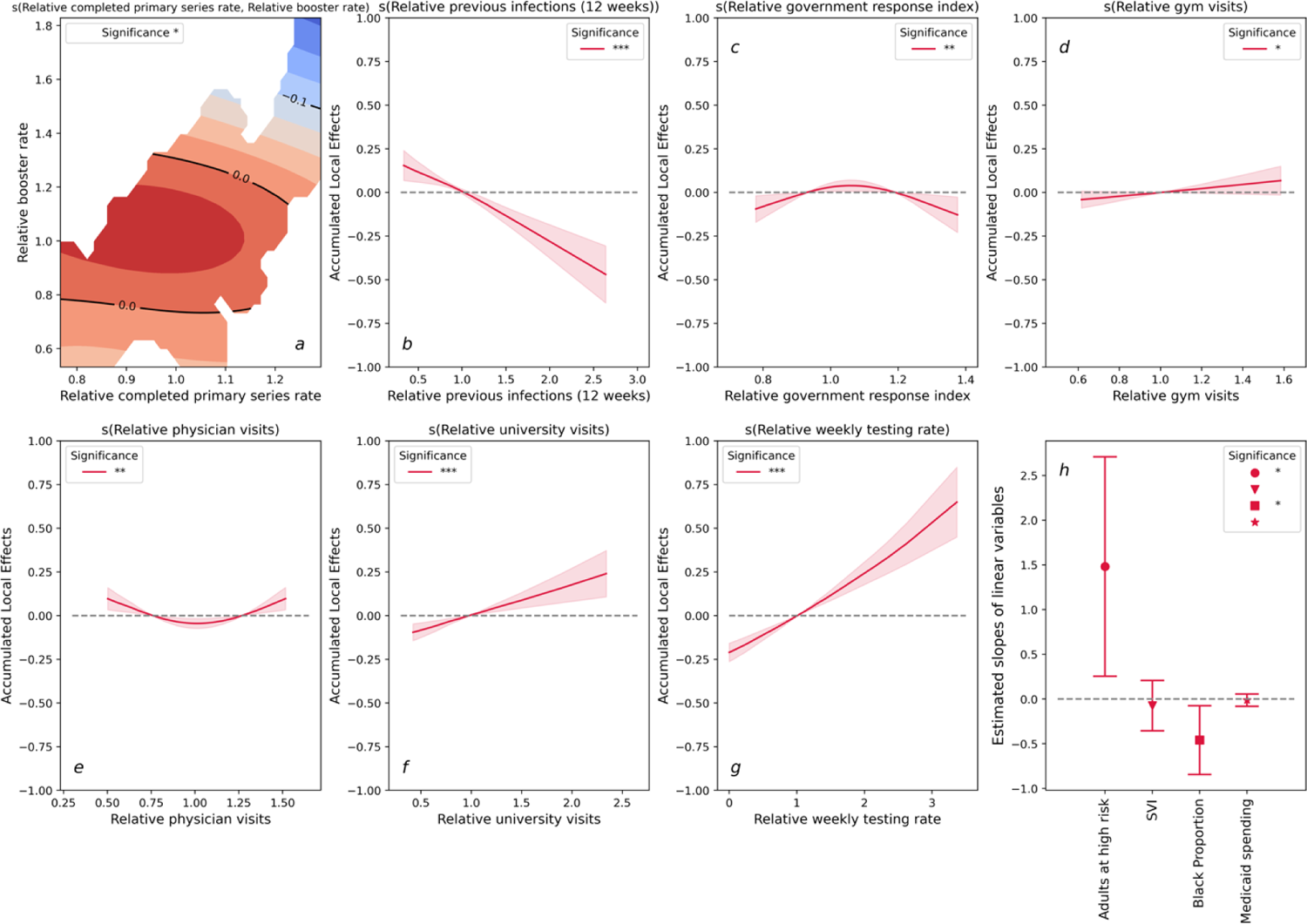
Results of the Omicron-Booster-RCIR for just the Omicron wave with the additional inclusion of an interaction effect between the relative completed primary series rate and the relative booster rate. a: Two-dimensional contour plot for the interaction between relative completed primary series rate and relative booster rate. b-g: Accumulated local effects (ALE) of dynamic variables. Shaded areas in each plot indicate 95% confidence intervals. h: Estimated slopes for each static variables, the upper and lower band indicate 95% condifence intervals. ‘***’: variable significant at p<0.001. ‘**’: variable significant at p<0.01. ‘*’: variable significant at p<0.05. ‘’: variable significant at p>0.05.

The incorporation of the relative booster rate does not result in an improvement in the model fit; the deviance explained for Model Omicron-Booster-RCIR remains at 17%. As depicted in Figure 4a, it is evident that only states with both a high relative completed primary series rate and a high relative booster rate exhibits a slightly negative impact, approximately −0.1, on the RCIR. The findings from Model Omicron-RCIR, when combined with Omicron-Booster-RCIR, suggest that the covariates examined in this study do not contribute significantly to explaining the variation in RCIR during the Omicron wave. These results highlight the need for further research to identify other factors that may better capture the dynamics of COVID-19 transmission during this specific period.

## 4. Discussion

This analysis aims to characterize the relationship between population-level COVID-19 vaccine administration and pandemic-induced healthcare burdens, taking into account essential and confounding real-world processes. Our results point to three significant conclusions:

- Population-level vaccination is always significantly associated with reduced COVID-19 case-hospitalization risk.
- Increased recent (1-4 months prior) infections are also consistently and strongly associated with reduced case-hospitalization risk.
- Local factors, activity-related engagement levels, and policy measures are important to the model’s explanatory power, supporting the importance of considering these factors on population-level outcomes. However, their associations are inconsistent over time and across different variants.

Each of these conclusions is explained in more detail in the sections below. In each section, we discuss the findings regarding case-hospitalization risk and compare them with the results related to the reported case incidence rate. In general, our results strongly support the importance of population-level vaccination and align with extant research on the role of acquired immunity in reducing severe outcomes. However, it should be noted that the case incidence rate has a reduced association with vaccination during the Omicron wave and much less consistently meaningful associations with previous infection rates. Additionally, our analysis reflects the complexity of the evolution of human behavior during the pandemic, given the dynamic role of activity-related engagement levels and policy. It also supports the recognition of the epidemiological vulnerability of socially and economically underserved communities.

### 4.1 Vaccines protect against COVID-19 case-hospitalization risk for pre-Delta, Delta and Omicron waves

Our study reveals a strong and statistically significant association between vaccine uptake rates and reduced COVID-19 case-hospitalization risk. This relationship was consistent across each of the variant waves modeled, and is consistent with earlier findings that vaccine protection against severe illnesses does not significantly wane in response to new variants. In contrast, when we modeled reported case-incidence rates as the response variable, we observed a decreasing effect of vaccines from the pre-Delta to the Omicron wave (Figure 3a). This outcome aligns with existing literature highlighting the rapid waning of the vaccines’ effectiveness against infection [33,34]. Nonetheless, while vaccines may offer reduced protection against infection, our results indicate that they continue to provide substantial protection against hospitalization risk and help alleviate the burden on healthcare systems. Additionally, although the value of booster shots for protection against severe cases of COVID-19 is still being studied [35], results from our analysis provide evidence supporting the effectiveness of booster doses against hospitalization risk caused by the Omicron variant (Figure 2). Conversely, the findings obtained from our Omicron-Booster-RCIR model reveal that the interaction between the booster and completed primary series rates has a relatively limited impact on Omicron infection (Figure 4). However, it is crucial to emphasize that despite the diminished effectiveness of mRNA boosters against Omicron infections, vaccines still serve the essential purpose of reducing the harm of COVID-19 in the face of emerging variants.

### 4.2 Immunity from recent infection protects against COVID-19 case-hospitalization risk upon reinfection

Higher past COVID-19 infection levels in a population are associated with a decrease in COVID-19 case-hospitalization risk, indicating immunity gained from infection can provide some protection against severe disease in the event of reinfection in the future, but only for a limited period of time. Our study utilized the total number of cases reported in a 12-week window, ranging from 4 to 16 weeks prior to the time period modeled, as a proxy for recently acquired immunity, and found a strong negative association between the previous infection rate and future case-hospitalization risk. These results were consistent across the different variant waves. This finding aligns with other case-control studies that found previous infections showed strong effectiveness against severe, critical, or fatal COVID-19 [6,36]. Our analysis indicates that prior infections from up to 6 months ahead are associated with decreased hospitalization risk, but 4 to 16 weeks has the strongest effect (see Appendix sections 2.4 and 2.5 for this sensitivity analysis). While the waning of natural immunity has been established in molecular and clinical research [37], our analysis provides additional insight at the population-level. In our models with case-incidence rate as the outcome variable, we found an insignificant association between previous infection and case-incidence rate during the pre-Delta and Delta waves. However, during the Omicron wave, there was a significant negative association (Figure 3b). This finding contrasts with existing literature that found, at the individual level, previous infection protected against infection pre-Omicron, but this effectiveness decreases substantially during the Omicron wave [38]. Nevertheless, at the population level, the number of infected individuals is considerably higher during the Omicron wave than earlier, while a smaller proportion remains susceptible. Consequently, the cumulative impact of previous infections becomes more pronounced. These results highlight that previous infections have a variable and inconsistent impact on reinfection at the individual and population levels.

### 4.3 Local factors contribute to variation in COVID-19 health outcomes

Existing clinical and statistical studies [14,16,17] have identified critical indicators for COVID-19 health outcomes including demographics, comorbidities, social vulnerability index (SVI), and healthcare expenditures. Results from our model using RCHR as outcome variables indicate that the SVI is positively associated with COVID-19 case-hospitalization risk across all variant waves (Figure 1i). This finding is consistent with existing literature [14,17], which suggests that individuals from socially vulnerable regions are more likely to experience harmful COVID-19 outcomes. For each new variant wave, the proportion of adults at high risk was less associated with case-hospitalization risk than for the prior wave (Figure 1h). This result aligns with a cohort study that the hazard ratio of hospital admissions with the Omicron variant, compared to the Delta variant, showed a more significant drop in the elder age group compared to individuals younger than 20 [39]. Our results reveal an insignificant association between black proportion and case-hospitalization risk during the Delta and Omicron waves, which differs from previously identified positive associations across all waves [14]. In the United States, the eligibility for Medicaid varies by state, but generally, individuals and families with incomes up to 138% of the federal poverty level may qualify for coverage [40]. Our results reveal a consistent negative association between state-level Medicaid spending per person and COVID-19 case-hospitalization risk (Figure 1k), which indicates the potential protective effect of healthcare expenditures in mitigating the impact of the pandemic on vulnerable groups. In contrast to the case-hospitalization risk models, the case-incidence rate models indicate that there is no evidence for consistent or significant associations with demographics, SVI, or healthcare expenditures across variant waves, except for adults at high risk consistently positively associated with case-incidence rate (Figure 3). These results suggest that dynamic COVID-19 infection risk is complex and changes over time, and the factors contributing to transmission vary across waves. Further research is needed for a more comprehensive understanding of the complex and evolving nature of COVID-19 transmission.

### 4.4 Activity-related engagement levels are associated with COVID-19 health outcomes

At the beginning of the pandemic, several studies evaluated the association between mobility and COVID-19 transmission with inconsistent findings [41,42]. One possible reason for this inconsistency is that aggregated mobility data may not accurately reflect the risk of dynamically changing human behaviors, given that a minority of travel activities could be accountable for a significant majority of infections [43]. Furthermore, the connection between mobility and harmful health outcomes remains unclear. Our study uses disaggregated mobility patterns to capture diverse behaviors between populations, specifically relative activity-related engagement levels, to explore the association between these variables and COVID-19 severity. To achieve this, we divided activities into subcategories based on their purpose. University visits were used to represent school-related activities, gym visits to signify high-risk indoor activities, and physician visits to indicate healthcare-related visits.

Results shown in Figure 1d indicate that the state-week pairs with relatively higher gym visits are expected to observe higher case-hospitalization risk during the Pre-Delta wave. However, this association did not reach statistical significance during the Delta and Omicron waves. In contrast, when we modeled the case-incidence rate as the outcome variable, our analysis revealed a minor effect of gym visits, as shown in Figure 3d. The positive impact of gym visits during the Pre-Delta wave may be linked to infections among unvaccinated individuals engaging in indoor activities. It is supported by existing research that unvaccinated individuals have a 2.6 times higher likelihood of contracting SARS-CoV-2 than vaccinated individuals during indoor activities [44]. Moreover, unvaccinated individuals exhibit a higher likelihood of hospitalization [45], leading to a strong positive association between gym visits and case-hospitalization risk during the initial phases of vaccination distribution. In addition to indoor activity, we also observed a significant association between case-hospitalization risk and overall visits to hospitals, medical centers, and Outpatient Care Centers. Unlike indoor activity, this association transitioned from negative to positive between the pre-Delta to Omicron wave. One possible explanation for this finding is that as the pandemic evolved, the public became more familiar with the disease and more tolerant of at-home symptom management; thus, those COVID-19 patients that sought medical care were more likely to be those with more severe symptoms. Finally, visits to the university were found to have a relatively minor impact on case-hospitalization risk. We hypothesize that this is due to the young and relatively healthy demographic that frequents visiting schools, while still vulnerable to contracting the SARS-CoV-2 virus, they are less likely to experience severe outcomes from COVID-19 infection. This hypothesis is further supported by the findings from the case-incidence rate model, which identified a positive association with university visits during the Delta and Omicron waves (Figure 3d). It is worth noting that during the Pre-Delta wave, school visits negatively impacted the case-hospitalization risk. However, this impact changed to a positive association for the later waves. These observations align with existing research, which has demonstrated that the younger population exhibits the highest increase in susceptibility to the Delta variant compared to the pre-Delta variant [46].

### 4.5 More stringent government public health policy is associated with reduced COVID-19 case-hospitalization risk

Our results indicate that more stringent government policies were associated with reduced COVID-19 case-hospitalization risk during the Pre-Delta and Delta wave. This is consistent with previous studies [47]. In particular, we found that state-week pairs with a significantly high government response index (indicating stricter policy) have a stronger negative effect on the case-hospitalization risk (Figure 1c). However, this negative effect decreased over time, and was least evident during the Omicron wave. The reduced effect of the policy during Omicron is likely due to a complex combination of factors, including the increasing population level immunity from both widespread adoptions of vaccines and prior exposure providing more protection from severe disease during this period, combined with a reduction in the government’s response to the pandemic over time.

Additionally, weekly testing rates were shown to be negatively associated with case-hospitalization risk. While this result does not imply a causative relationship between testing rate and COVID-19 severity, there are various reasons why testing rates may be linked to case-hospitalization risk. Firstly, it represents a proxy input feature to capture the level of healthcare infrastructure available to a population. Second, it directly impacts the reported case incidence rate, as the number of reported cases in a region is a direct function of local testing availability, thus increased testing will lead to higher reported case rates, and lower case-hospitalization risk. Third, increased testing can lead to more cases being identified, and thus impact people’s awareness and behavior during an outbreak. For these reasons testing rate is included as a potential control factor in our model.

### 4.6 Limitations

As with all modeling studies, this work is subject to several limitations. Firstly, this study was primarily designed to determine the association between various potential risk factors and COVID-19 outcomes, rather than to establish causality between these variables. Thus, our findings may reflect the role of unobserved confounding factors excluded from our study. Another potential limitation is due to the application at the state-level. The aggregation of the data to the state-level is unable to capture the heterogeneities of the communities within each state, and it is possible that different associations exist at the local level, than are identified at the state-level. Additionally, while we believe the use of the case-hospitalization risk in a given state at a given time is a plausible choice as a proxy for disease severity at an individual-level, and captures the burden on the healthcare system at a population-level. However, it is subject to variable case reporting and data quality across states, which may arise due to uneven testing capacity, reporting delays or at-home testing. Lastly, it is important to acknowledge that our variable transformation, while facilitating a deeper understanding of relative changes, does come with the inherent consequence of diminishing the original meaning these variables initially conveyed.

## 5. Conclusions

This research utilizes publicly available real-world data to provide robust evidence of the efficacy of vaccines against COVID-19 case-hospitalization risk across various variant waves in the United States. More importantly, this paper concludes that booster shots offer additional protection against severe COVID-19 during the Omicron waves. Despite the emergence of new variants, vaccines remain the most effective intervention for mitigating the harm of COVID-19 and reducing burden on healthcare systems. Therefore, given the ongoing threat posed by COVID-19 and its potential variants, vaccines continue to be the best line of defense for protecting public health and preventing the further spread of the virus.

## Availability of data and materials

All the data and code used for the analysis is available from https://github.com/hongru94/vaccination_rate_GAMs.

## Author’s Contributions

LG, and HD contributed to the conceptualization and design of the study. HD and SS collected the data and conducted the analysis. HD led the writing of the original draft. HD, SS, and LG edited the manuscript, discussed the results, and provided feedback regarding the manuscript. LG supervised the study and acquired funding. HD and SS have verified the underlying data. All authors had full access to the data and approved the manuscript for publication.

## Data Availability

All data produced are available online at https://github.com/hongru94/vaccination_rate_GAMs

## Acknowledgments

We want to thank Maximilian Marshall for providing valuable insights and suggestions that have contributed to enhancing the quality of this paper.

## Supplementary Materials for

## 1. Supplementary Data

### 1.1 Preprocessing of genomic data

All genomic data were collected from GISAID [1] on October 27^th^, 2022. GISAID is a publicly accessible repositor of dataset that sharing of genomic data on various pathogens, including influenza and COVID-19. We analyzed the available set of sequences, to determine the proportion of each variant theoretically in circulation. Specifically, we calculated the proportion of each variant for each week in each state from March 1^st^, 2021, to March 1^st^, 2022. T identify the most dominant variant for each state-week pair during the analyzed period, we labeled the state-week pairs based on the variant with the highest proportion. This enables us to track the dominant variant in each state an cluster the state-week pairs based on the most dominant variant. The assignment of state-week pairs is shown in Appendix figure S1 below:

**Appendix figure S1:**
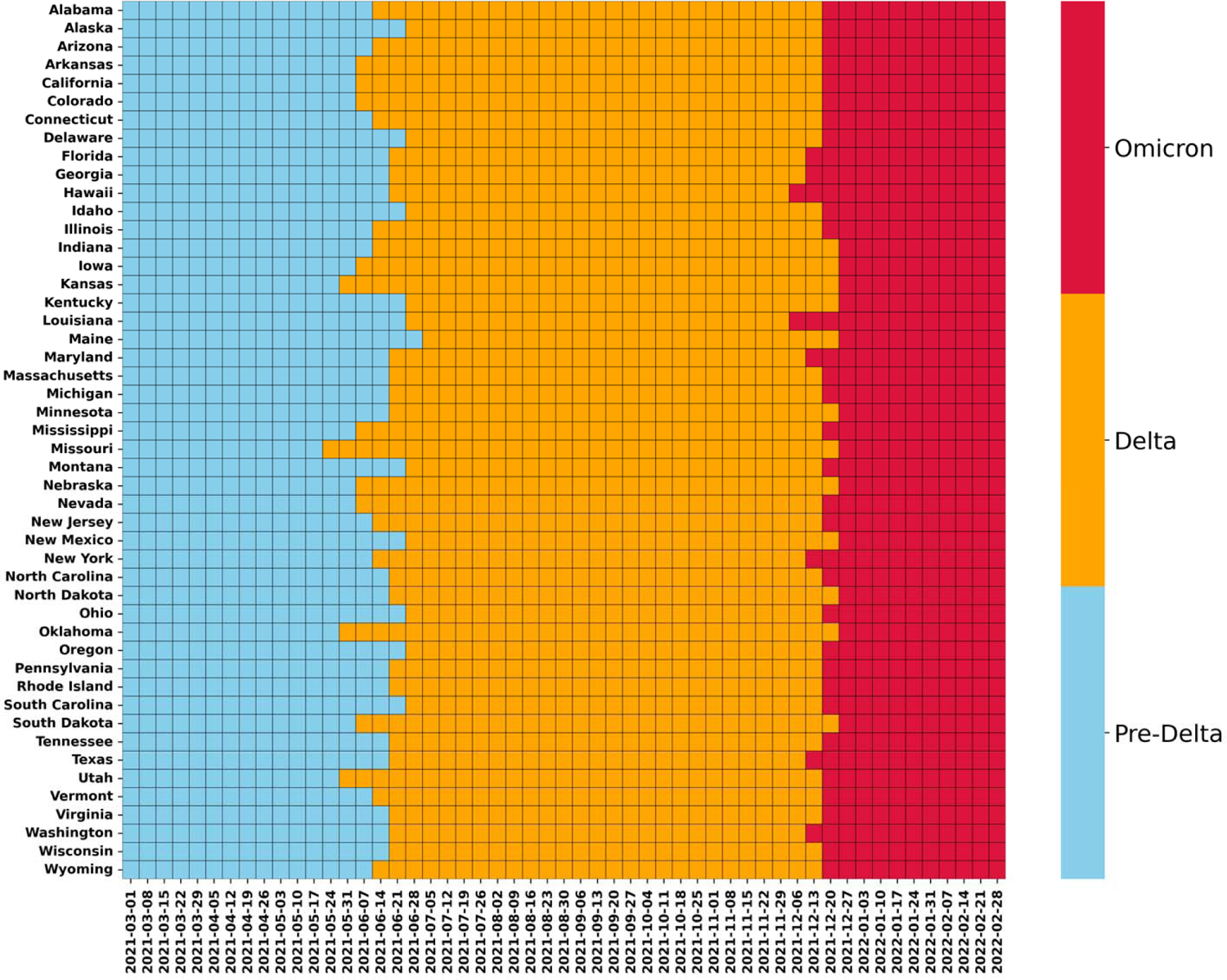
State-week group assignment based on the dominant variant. The x-axis represents each week, the y-axis represents each state, and the color represents the assignment of each state-week pair.

## 1.2 Variables description

### Outcome variable

Case-hospitalization risk: In this study we used case-hospitalization risk (CHR_i_^t^) as the outcome variable of interest for our model. COVID-19 case-hospitalization risk represent both the severity of COVID-19 disease at an individual level and the burden it places on the healthcare system. Case-hospitalization risk (CHR_i_^t^) for each state *i* and week *t* is defined as follow:

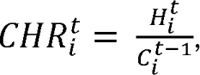

where *H_i_^t^* is the number of hospitalizations for state *i* and week *t* and C_i_^t-1^ is the number of confirmed cases for state *i* and week t-1. We applied a one-week lag between reported confirmed cases and hospitalizations to account for the time between symptom onset and hospital admission [2]. Weekly state-level CHR^t^ was treated as the outcome variable in this analysis and served as an indicator for the burden of COVID-19 risk for a given time and location.

### Dynamic Covariates

Vaccination rate: In this analysis, the weekly cumulative COVID-19 vaccination rate is the primary variable under examination, as we hypothesize it to be a critical determinant in protecting populations against severe COVID-19 disease. The completed primary series rate was chosen over the partial vaccination rate because it represents the recommended dosage by the U.S. CDC. To address the fact that vaccine eligibility was not available to all U.S. adults until April 19^th^, 2021, we also ran the model using the partial vaccination rate for the pre-Delta waves, and the results are consistent with the completed primary series rate (results are presented in Appendix section 2.3). As booster shots became widely available during the Omicron wave, we also include the booster vaccination rate as a covariate to investigate its potential impact on COVID-19 health outcomes. Due to errors and anomalies in the data, the vaccination data excludes West Virginia and New Hampshire, while the booster vaccination data excludes West Virginia, New Hampshire, and North Carolina. As a result, West Virginia and New Hampshire are excluded from all analyses, and North Carolina is excluded from the booster analysis for the Omicron wave.

Activity-related engagement level: We adapted multiple mobility-derived metrics from a previous study [3] to represent destination-specific travel behaviors and activity-related engagement levels for specific types of activities, namely gym, university, and physician visits. Specifically, the engagement levels represent the weekly number of visits to a given destination of interest per person per week. This variable allows us to compare the relative frequency of visits to each point of interest across states and to investigate their potential impact on COVID-19 health outcomes. The metrics were generated based on anonymized mobility data from Safegraph [4], which tracks the number of visits to different types of destinations for a sample of the population at the census tract level in the U.S. Examples of destinations include full-service restaurants, gyms, and grocery stores. The original Safegraph dataset includes over 20 destination categories; thus, to reduce the complexity of the model we identified a smaller representative set of destinations to include as input in the final model. This was accomplished by first organizing the destination categories into six distinct destination groups based on the first two digits of the NAICS code [5], namely Retail Trade (44-45), Education Services (61), Healthcare and Social Assistance (62), Arts, Entertainment, and Recreation (71), Accommodation and Food Services (72), and Other Services (81). From each group, we selected one destination category as the representative variable for the group based on the correlations between other variables within the group (Details are documented in Appendix section 1.4). Subsequently, we conducted a model selection process to identify the most appropriate subset of mobility variables from these six to be included in the final model based on concurvity and significane level (details are documented in Appendix section 2.2).

Previous infection rates: Several studies have demonstrated the effectiveness of previous infections against reinfections and severe COVID-19 outcomes. Studies have illustrated that individuals retain a substantial level of natural immunity for six months after infection [6–8]. To attempt to account for the role of recently acquired immunity from infection in protecting from severe disease upon reinfection in our study we generate a variable to represent the total population infected and recovered within a recent window, i.e., the total infections reported between weeks (t-16) and (t-4), which allows for time to recover and build up immunity [9] by the time period t at which the hospitalizations are modeled, but short enough that immunity has not waned. These specific prior infections variable (PI^t^), requires multiple parameters, namely the length of the interval that infections are summed over and the start and end period of the window. To identify the best window and evaluate the sensitivity of our analysis to the chosen window length, start and end time, we conducted a sensitivity analysis. The time window with the largest deviance explained in the GAMs was selected for the final model, which was a three-month window ranging from 4 to 16 weeks prior to time t. Additional details of this sensitivity analysis and the results are included in Appendix section 2.4. The mathematical formulation of this metric is defined as follows:

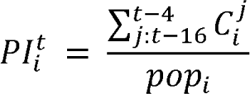

where *PI_i_^t^* represents cumulative infection rate for state *i* from 16 to 4 weeks prior of week *t, C_i_^j^* is the weekly confirmed cases for state *i* at week *j*, and pop_l_ is the population for state i. The sum in the numerator defines the summation of C^j^ for the t-16 to t-4 weeks prior to t.

Government policy: The stringency and timing of implementing government policies to mitigate the impacts of COVID-19, such as school closure, cancellation of public events, and international travel controls, are associated with different measures of epidemic severity [10]. We selected the government response index from Oxford Coronavirus Government Response Tracker (OxCGRT) [11] as our indicator for government policy. The index tracks the diversity of government responses across various policies, ranging from containment measures and closures to healthcare systems, vaccination strategies, and economic policies. This index reflects the government’s response level with a number ranging from 0 to 100, the larger the number, the more substantial the response. It is available for all 50 states in the U.S. at a weekly timescale for the entire period of analysis.

Weekly testing rate: The weekly testing rates were included in the model as a potential confounding factor for multiple reasons. Firstly, it represents a proxy input feature to capture the level of healthcare infrastructure available to a population. Second, it directly impacts the case-hospitalization risk through the denominator (i.e., total reported cases), as the number of reported cases in a region is a direct function of local testing availability, thus increased testing will lead to higher reported case rates, and lower case-hospitalization risk. For example, in two locations with the same true case-hospitalization risk (e.g., the likelihood of a COVID-19 infection needing admittance is equal), a location with twice as much testing will detect more cases, and therefore appear to have a lower case-hospitalization risk. Third, increased testing can lead to more cases being identified, and thus impact people’s awareness and behavior during an outbreak. For these reasons testing rate is included as a potential confounding factor in our model. We normalized the raw weekly total testing count by population to get the weekly testing rate.

**Appendix figure S2:**
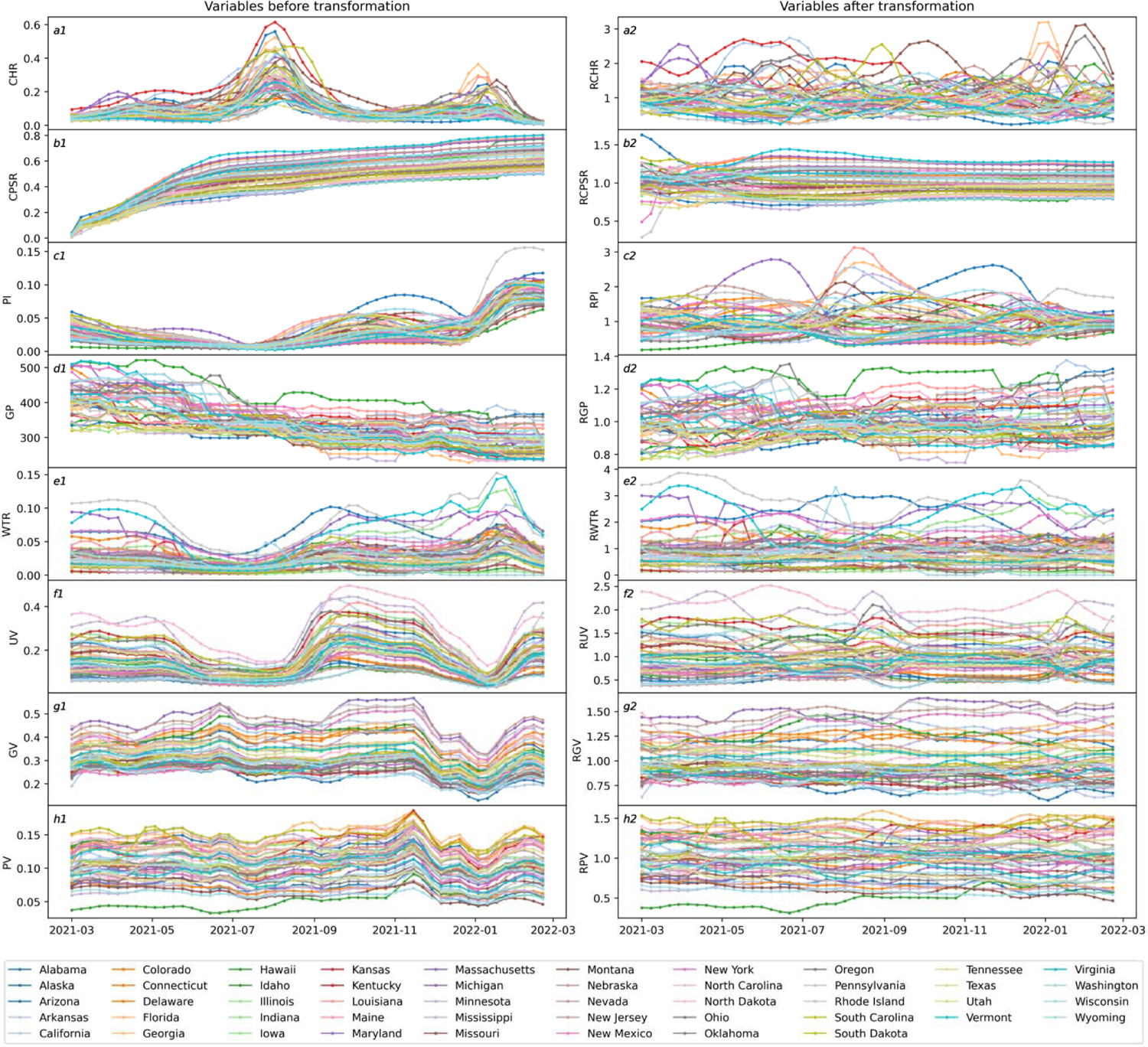
Visualization of dynamic variales before variable transformation (a1 to h1) and after variable transformation (a2 to h2). For the y-axis label, the abbreviations signify the following variables: CHR: Case-hospitalization risk, CPSR: Completed primary series rate, PI: Previous infection, GR: Government policy measure, WTR: Weekly testing rate, UV: University visits, GV: Gym visits, PV: Physician visits.

## Static Covariates

**Appendix table S1:** Full list of static variables.

| Variable name | Variable description | source |
| --- | --- | --- |
| Static variables |  |  |
| Black proportion | The proportion of the population identified as non-Hispanic Blacks. | [13] |
| Medicaid spending | Total Medicaid spending in kilo for each state normalized by the population. | [14] |
| Healthcare spending | Total Healthcare spending in kilo for each state normalized by the population. | [15] |
| Poverty rate | Percentage of population living below poverty line. | [16] |
| Social Vulnerability Index | The Social Vulnerability Index utilizes data from the U.S. Census to assess the relative level of social vulnerability in each census tract. By analyzing 14 social factors, the SVI categorizes tracts into four closely interrelated themes and then aggregates them as a single indicator of social vulnerability. | [17] |
| HAQI | IHME's healthcare access and quality index. | [18] |
| Republican voters | Percentage of a state's voters who voted for the 2020 Republican presidential candidate. | [19] |
| Adults at high risk | The proportion of the population over 18 years old is at high risk of serious illness if infected with Coronavirus. | [20] |
| Proportion over 65 | Proportion of population age 65 and older. | [20] |

### 1.3 Preprocessing of mobility data

The 21 mobility destination categories from Safegraph were organized into six distinct industry groups based on the NAICS code. The relevant groups per the NAICS code are Retail Trade (44-45), Education Services (61), Healthcare and Social Assistance (62), Arts, Entertainment, and Recreation (71), Accommodation and Food Services (72), and Other Services (81).^2^ The details of generating visits to each POI are documented in previous publication.

**Appendix table S2:** Description of each industry group and the corresponding destination categories.

| Retail Trade (44-45) | Education Services (61) | Healthcare and Social Assistance (62) | Arts, Entertainment, and Recreation (71) | Accommodation and Food Services (72) | Other Services (81) |
| --- | --- | --- | --- | --- | --- |
| Automotive Store (441310) | Elementary School (611110) | Office of Physician (621111) | Parks (712190) | Hotels (721110) | Religious Organizations (813100) |
| Hardware Store (444130) | University (611310) | Child Day Care (624410) | Gym (713940) | Full-Service Restaurant (722511) |  |
| Grocery Store (445110) |  |  |  | Cafes, Snacks, Bars (722515) |  |
| Convenience Store (445120) |  |  |  | Limited-Service Restaurant (722513) |  |
| Pharmacies (446110) |  |  |  |  |  |
| Gas Station (447110) |  |  |  |  |  |
| Sporting Goods Store (451110) |  |  |  |  |  |
| Department Store (452210) |  |  |  |  |  |
| Other General Store (452319) |  |  |  |  |  |
| Used Merchandise Store (453310) |  |  |  |  |  |

To reduce the complexity of the model, we selected one destination category as the representative variable for each industry group. For the industry groups with more than 3 destination categories, we conducted a Pearson’s correlation analysis and selected the variable that had the highest correlation to the other destination categories in each group. This method selected Gas Stations and Full-Service Restaurant from the Retail Trade (44-45) and Accommodation and Food Services (72) as the representative variable for each industry group. For the Educational Services (61) and Healthcare and Social Assistance (62) groups, we selected University and Office of Physician as the representative variables based on studies that indicated SARS-CoV-2 infection severity is lower in adolescents than adults.^3^ For the Arts, Entertainment, and Recreation (71) industry group, we selected Gym as the representative variable instead of Parks because studies have identified park use to have a minor effect on COVID-19 transmission compared to other mobility destinations.^4^ Religious Organizations was selected from the Other Services (81) industry group because it is the only destination category present.

## 2. Supplementary Methods

### 2.1 Static variables selection

We selected state-level static variables that were found to have association with COVID-19 health outcomes in a recent study [12]. These variables cover a ranging of different factors, such as socioeconomic indicators, racial demographics, age, proxy for comorbidities, political factors, and state-level healthcare expenditures. Then, a correlation analysis is performed within static variables to determine the suitable variables to be included in the model. Full list of static variables included in the correlation analysis are listed in Appendix table S1.

**Appendix figure S3.**
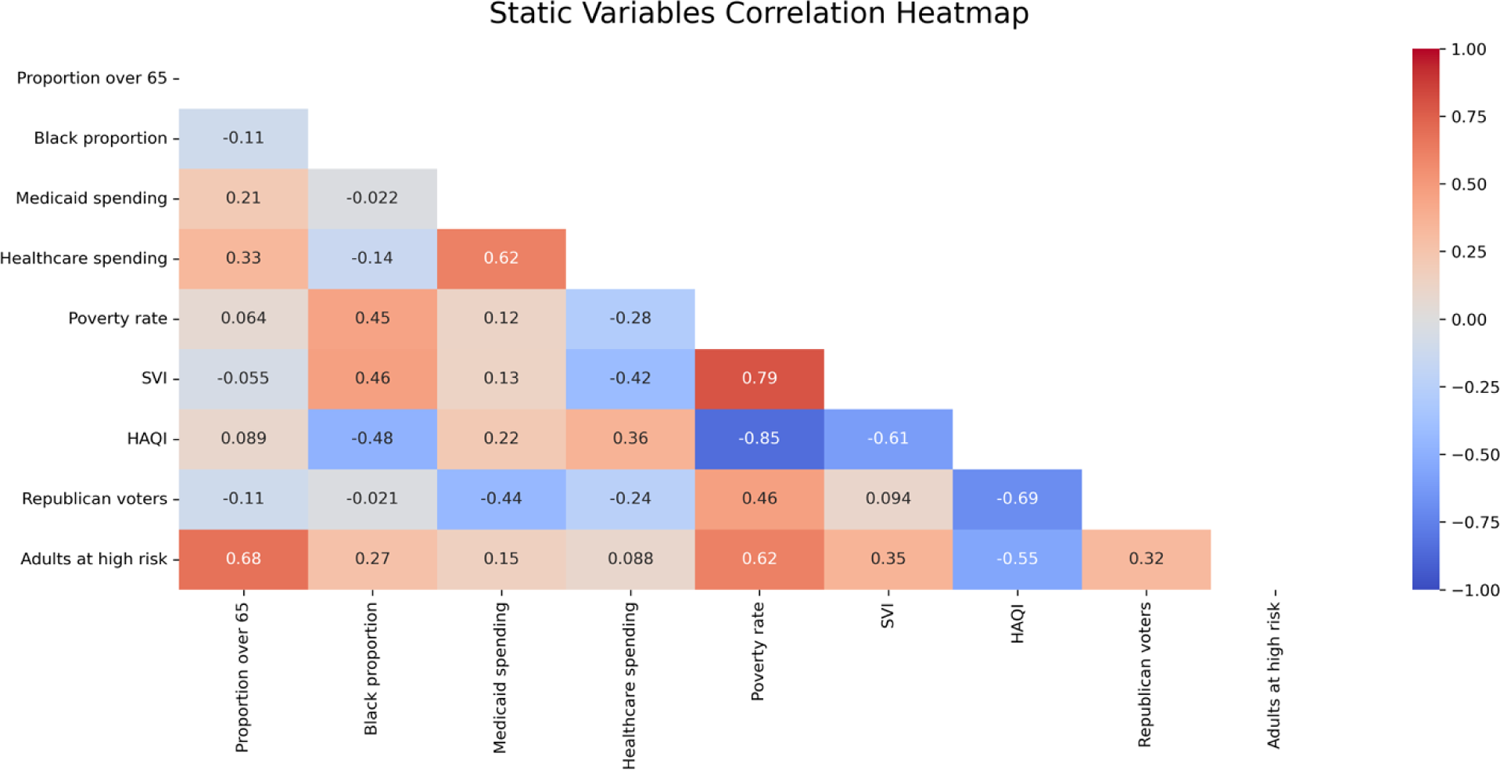
Pearson’s correlation heatmap between each pair of static variables.

Based on the correlation analysis, the set of static variables to be incorporated into the models was determined. We selected the black proportion as a representative variable for the race group and the SVI as a proxy for the vulnerable population. The poverty rate was dropped due to its high correlation with SVI. Additionally, we selecte adults at high risk as a control variable for population-level comorbidities and dropped the proportion over 65. We also decided to include Medicaid spending over healthcare spending as the state-level proxy for healthcare expenditures. Lastly, we dropped the HAQI and Republican voters variables due to their high correlation with the completed primary series rate.

### 2.2 Dynamic variables selection

The GAMs fit outcome variables with smoothed independent variables, allowing the nonlinear relationships between input and output. However, the nonlinear variables smoothing sometimes can result in concurvity issues. Concurvity occurs when some smooth term in a model could be approximated by one or more of the other smoot terms, leading to inaccurate estimates of the effect for given variables. In this section, we conduct model selection *t* ensure the validity of our model and to detect and mitigate any concurvity issues that may arise, using mobility data selected from Appendix section 1.4 and other independent variables. The significance level and the concurvity for each variable for every model are reported in table S2 below:

**Appendix table S3:** Significance level and concurvity for all dynamic variables.

| Variable | Pre-Delta wave |  | Delta wave |  | Omicron wave |  |
| --- | --- | --- | --- | --- | --- | --- |
|  | Signif | Concurvity | Signif | Concurvity | Signif | Concurvity |
| Relative completed primary series rate | *** | 0.71 | *** | 0.79 | *** | 0.81 |
| Relative previous infection rate (12 weeks) | *** | 0.62 | *** | 0.31 | *** | 0.41 |
| Relative full-service restaurant rate |  | 0.82 |  | 0.81 | * | 0.86 |
| Relative gas station visits |  | 0.91 |  | 0.85 | ** | 0.89 |
| Relative religious organization visits | *** | 0.84 |  | 0.84 | ** | 0.87 |
| Relative gym visits | *** | 0.69 | ** | 0.76 |  | 0.76 |
| Relative university visits | * | 0.80 | *** | 0.62 | *** | 0.64 |
| Relative office of physician visits | *** | 0.70 |  | 0.65 | ** | 0.68 |
| Relative weekly testing rate | *** | 0.81 | *** | 0.64 | *** | 0.57 |
| Relative government response index | *** | 0.78 | *** | 0.32 |  | 0.27 |

Significance codes: '\*\*\*': 0.001, '\*\*': 0.01, '\*': 0.05, '.': 0.1, ':': > 0.1.

The value of concurvity range from 0 to 1, the higher the concurvity the more a smooth variable can be approximated by the smooth of other variables. Specifically, a concurvity value above 0.8 generally signals the need for careful inspection of the model. Based on the results form table S3, we removed relative full-service restaurant, relative gas station, and relative religious organization visits from the model. This decision was based on their lack of significance and/or their high concurvity values. The equivalent results for selected variables are presented in table S3.

**Appendix table S4:** Significance level and concurvity for selected variables.

| Variable | Pre-Delta wave |  | Delta wave |  | Omicron wave |  |
| --- | --- | --- | --- | --- | --- | --- |
|  | Signif | Concurvity | Signif | Concurvity | Signif | Concurvity |
| Relative completed primary series rate | *** | 0.63 | *** | 0.61 | *** | 0.60 |
| Relative previous infection rate (12 weeks) | *** | 0.59 | *** | 0.25 | *** | 0.35 |
| Relative gym visits | *** | 0.54 |  | 0.57 |  | 0.56 |
| Relative university visits |  | 0.69 | *** | 0.49 | *** | 0.44 |
| Relative office of physician visits | *** | 0.66 |  | 0.60 | ** | 0.64 |
| Relative weekly testing rate | *** | 0.78 | *** | 0.51 | *** | 0.51 |
| Relative government response index | *** | 0.72 | *** | 0.26 | * | 0.23 |

Significance codes: '\*\*\*': 0.001, '\*\*': 0.01, '\*': 0.05, '.': 0.1, ':': > 0.1.
The results from table S4 reveal that each variable exhibits a concurvity value below 0.8 and is significant in at least one out of three models.

### 2.3 Robustness check of vaccination data

We selected the completed primary series rate as the main vaccination variable in the main analysis. To assess the robustness and validity of our findings, we conducted additional analyses using different vaccination data (completed primary series rate, and partial vaccination rate) and varying starting dates (March 8^th^, 2021, and April 19^th^, 2021) for the analysis. We applied our sensitivity analysis to Model Pre-Delta-RCHR, as it is the only ones that could be affected by the analysis. The results of four different combination of vaccination data and starting date for Model Pre-Delta-RCHR are shown in Appendix figure S4 below:

The finding from this robustness check demonstrated a strong and consistent impact of vaccination, independent of the chosen vaccination data or the starting date of the analysis. This consistency suggests the robustness of our results and highlights the robustness of the completed primary series rate as the main vaccination variable.

### 2.4 Sensitivity analysis of prior window length for previous infection

In this section, we presented a sensitivity analysis to assess the impact of the prior window length for the previous infection on our analysis. To ensure our results are robust, we fixed all other covariates and a lag of four weeks for previous infections while varying the prior window length for previous infections from 12 to 24 weeks. The results of this sensitivity analysis for each model are shown below:

**Appendix figure S4:**
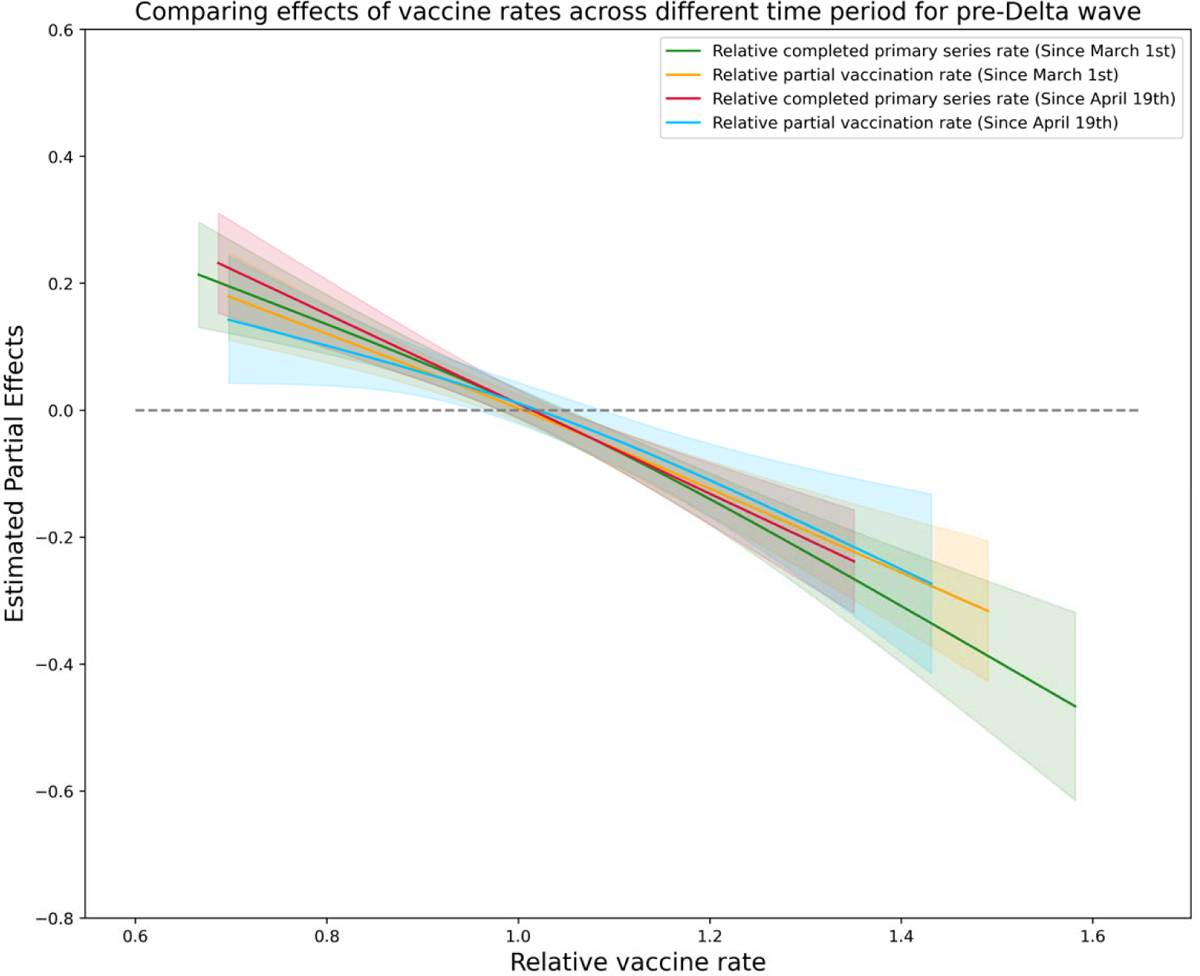
Robustness check of vaccination rate with different vaccination data and varying starting date for Model Pre-Delta-RCHR.

### 2.5 Sensitivity analysis of lags for previous infection

This section presents a sensitivity analysis to assess the impact of the prior window length for the previous infection on our analysis. To ensure our results are robust, we fixed all other covariates and a prior window length of 12 weeks for previous infections while varying the lag for previous infections from 4 to 16 weeks. The results of this sensitivity analysis for each model are shown below:

**Appendix figure S5:**
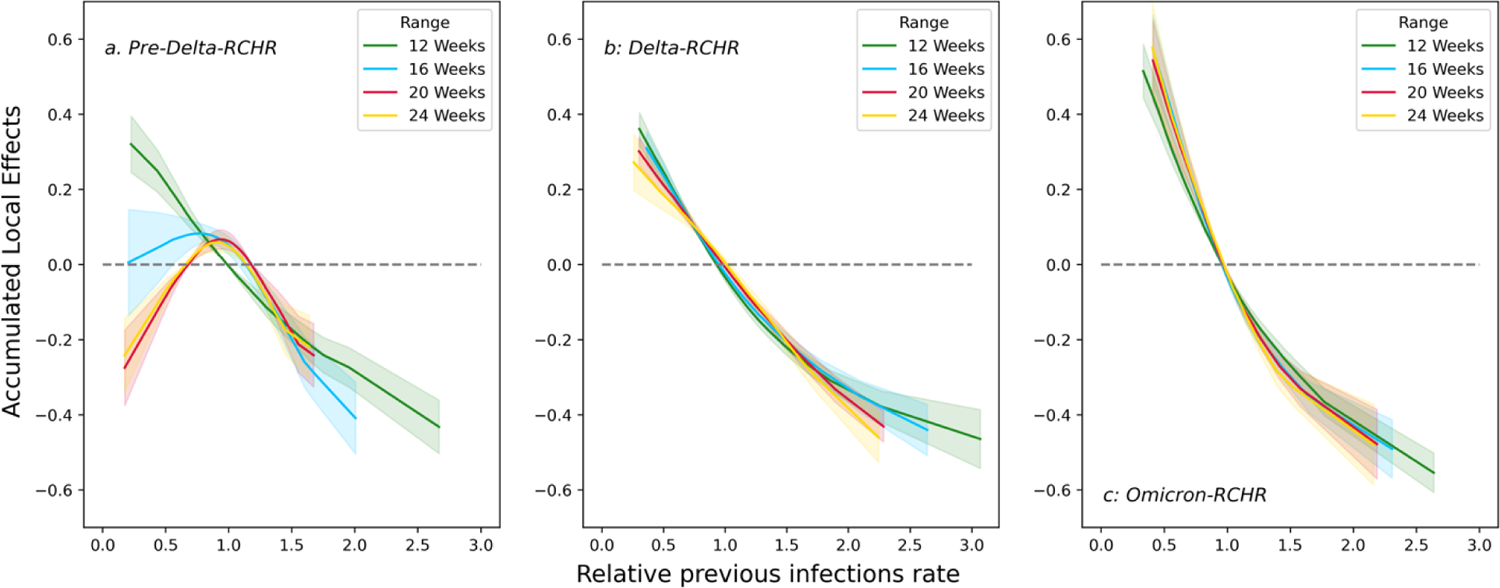
Comparison of accumulated local effects of the previous infection rate for Model Pre-Delta-RCHR (a), Delta-RCHR (b), and Omicron-RCHR (c) with different prior window lengths (12, 16, 20 and 24 weeks).

### 2.6 GAMs with reported case-incidence rate (RCIR) as the outcome variable

These GAMs share the same framework as Model Pre-Delta-RCHR, Delta-RCHR, and Omicron-RCHR, while the outcome variable is the reported case-incidence rate (RCIR). To account for the sequential process leading to infections, all lags between dynamic covariates and RCIR have been reduced by one week. These three GAMs have the form:

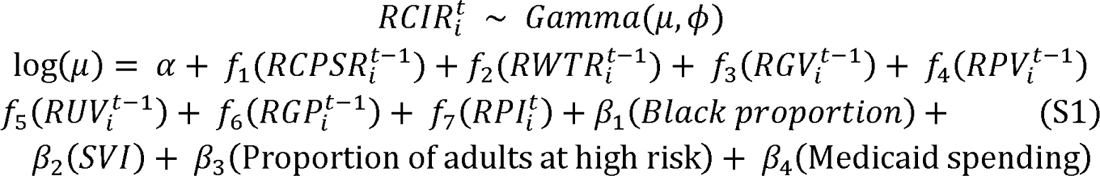

**Appendix figure S6:**
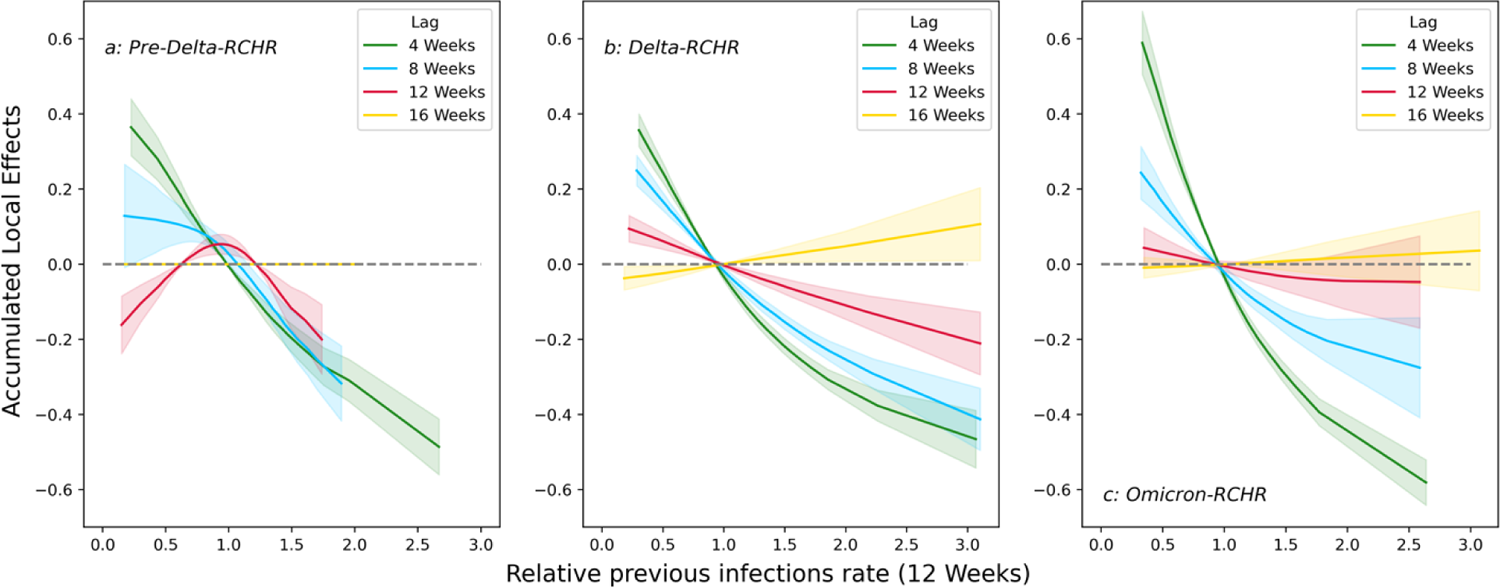
Comparison of the accumulated local effects of the previous infection rate for Model Pre-Delta-RCHR (a), Delta-RCHR (b) and Omicron-RCHR (c) with different lags (4, 8, 12, and 16 weeks).

Where α represents the intercept, β_l_ represent the parametric coefficients of each static variable, and *f_i_* are spline smooth functions of the relative dynamic variables. Additionally, a model is constructed for the Omicron wave, incorporating an interaction between completed primary series and booster rate (Omicron-Booster-RCIR). The model Omicron-Booster-RCIR has the form:

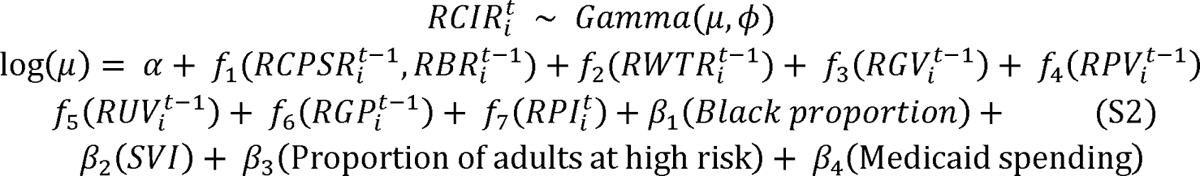

Where *f_1_* represent a smooth interaction function between RCPSR_i_^t-2^ and RBR_i_^t-2^. For all the mentioned models above, the weekly state-level RCHR is assumed to follow a Gamma distribution with a log link. This choice of the Gamma family accounts for the positively skewed distribution of the outcome variable. We use thin plate regression splines as the smoothing basis for all *f_i_* and set the basis dimension to three to maximize the interpretability of the models.

## 3. Supplementary Results

### 3.1 Models evaluation for GAMs with RCHR as outcome variable

**Appendix figure S7:**
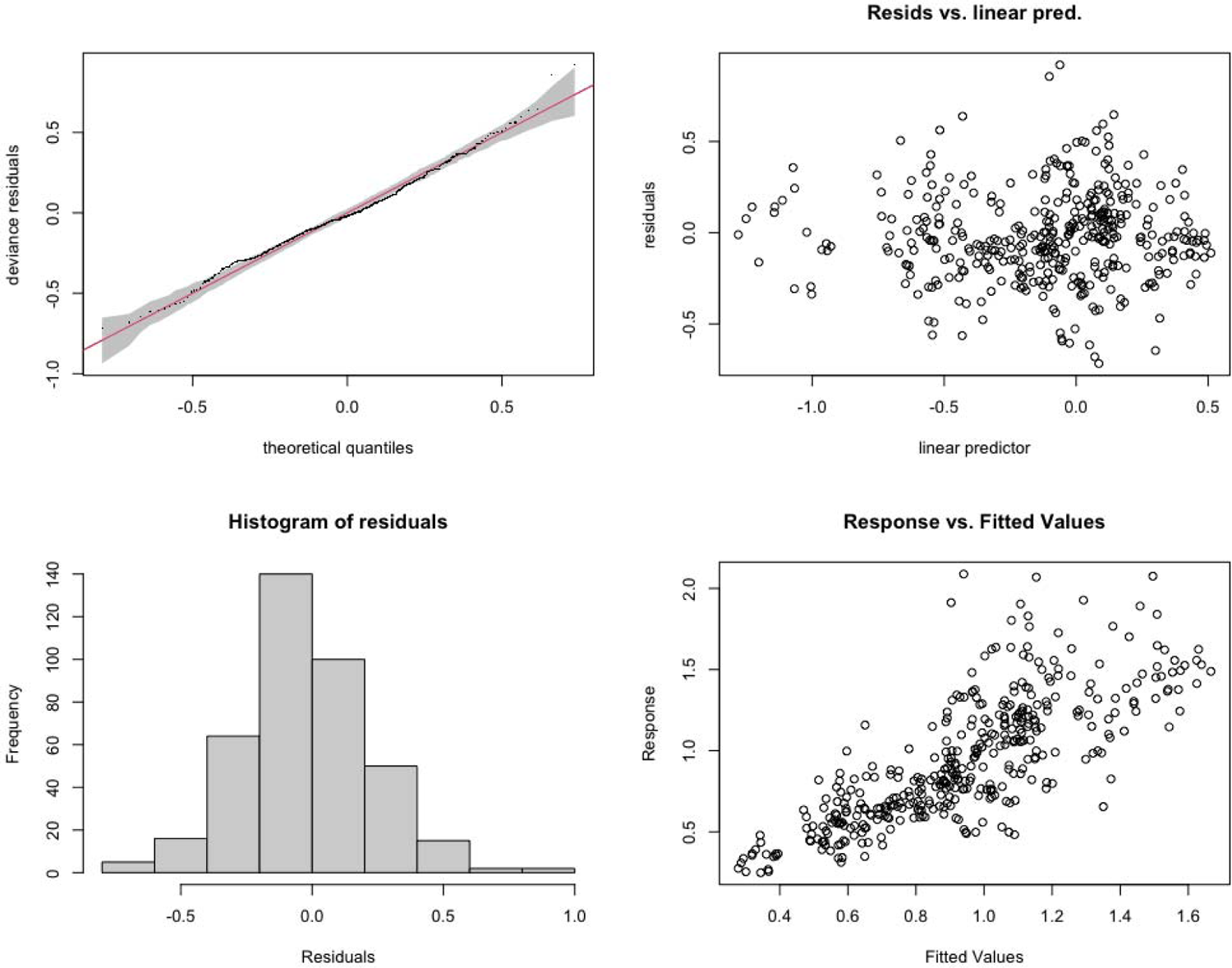
Model diagnostic plots for Model Pre-Delta-RCHR. The correlation coefficient between fitted RCHR and predicted RCHR is 0.78.

**Appendix figure S8:**
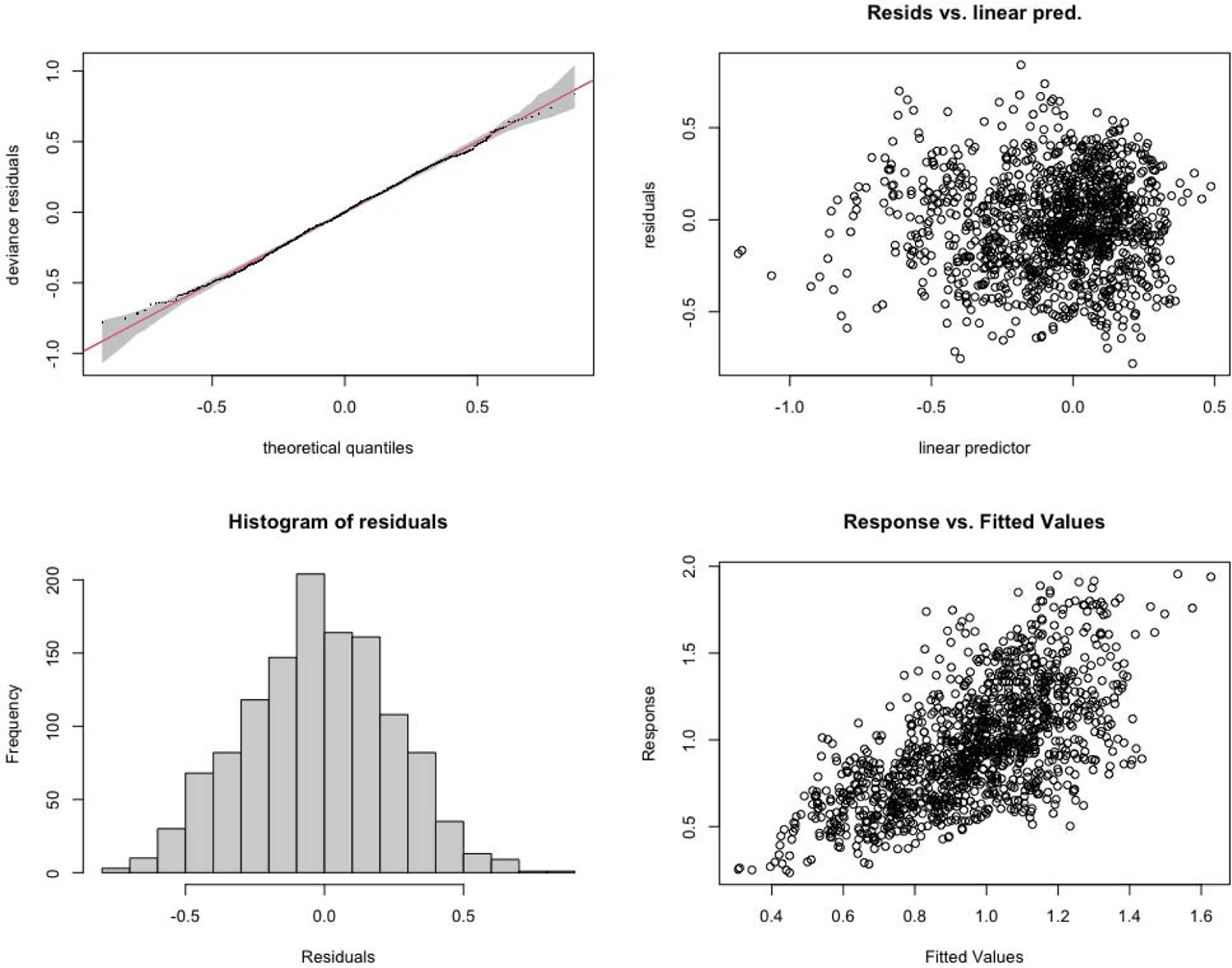
Model diagnostic plots for Model Delta-RCHR. The correlation coefficient between fitted RCHR and predicted RCHR is 0.67.

**Appendix figure S9:**
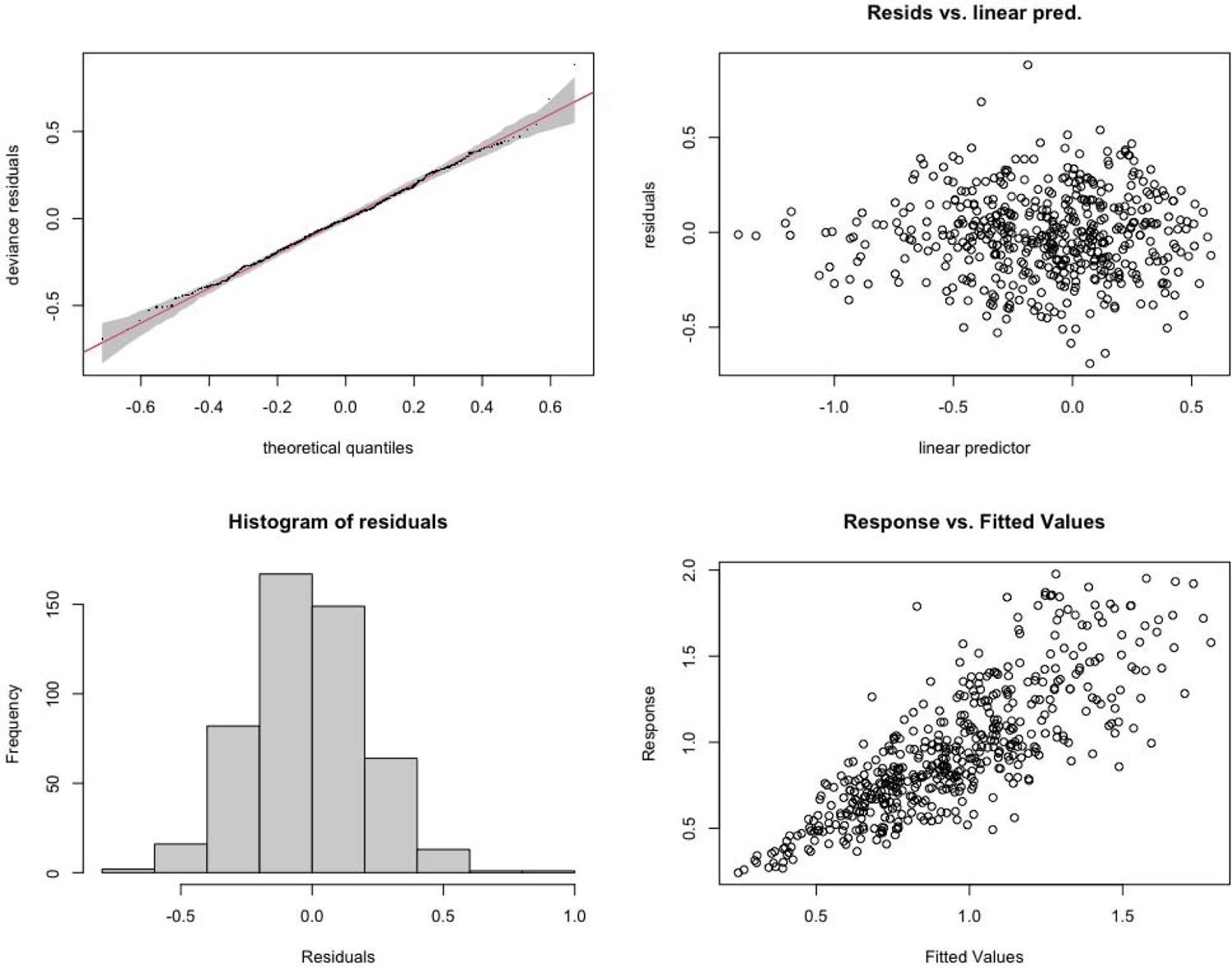
Model diagnostic plots for Model Omicron-RCHR. The correlation coefficient between fitted RCHR and predicted RCHR is 0.81.

**Appendix figure S10:**
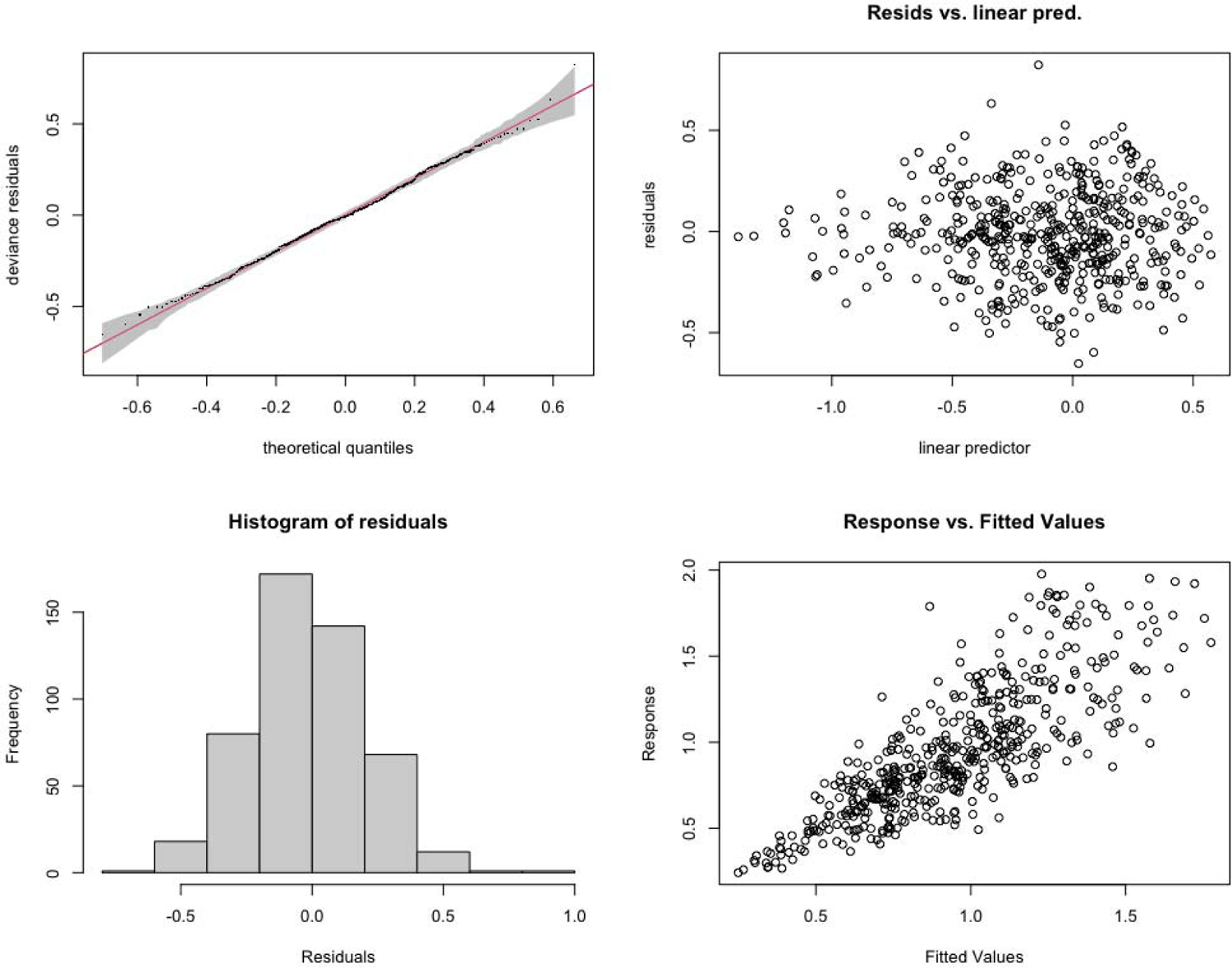
Model diagnostic plots for Model Omicron-Booster-RCHR. The correlation coefficient between fitted RCHR and predicted RCHR is 0.83.

### 3.2 Models evaluation for GAMs with RCIR as outcome variable

**Appendix figure S11:**
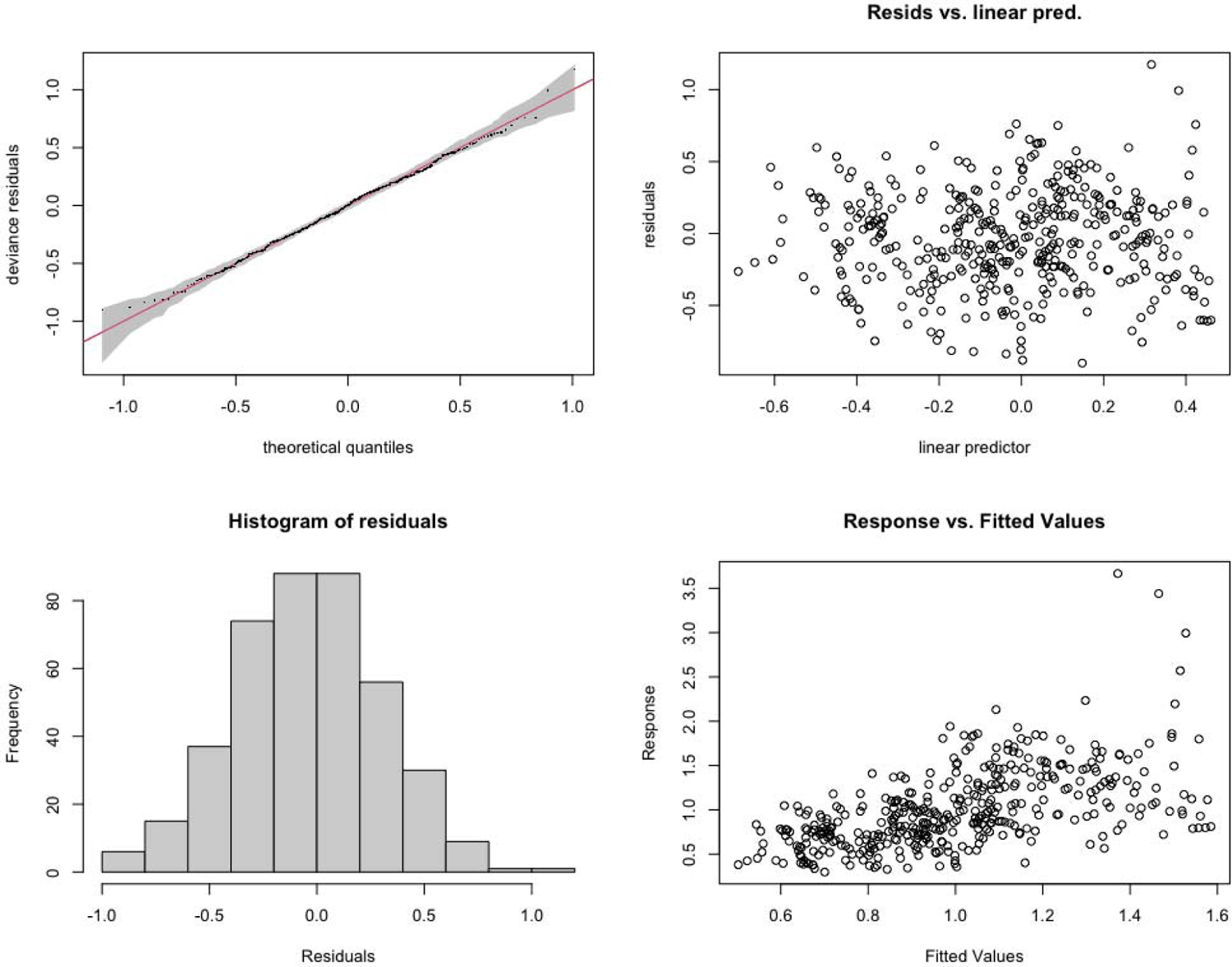
Model diagnostic plots for Model Pre-Delta-RCIR. The correlation coefficient between fitted RCHR and predicted RCHR is 0.57.

**Appendix figure S12:**
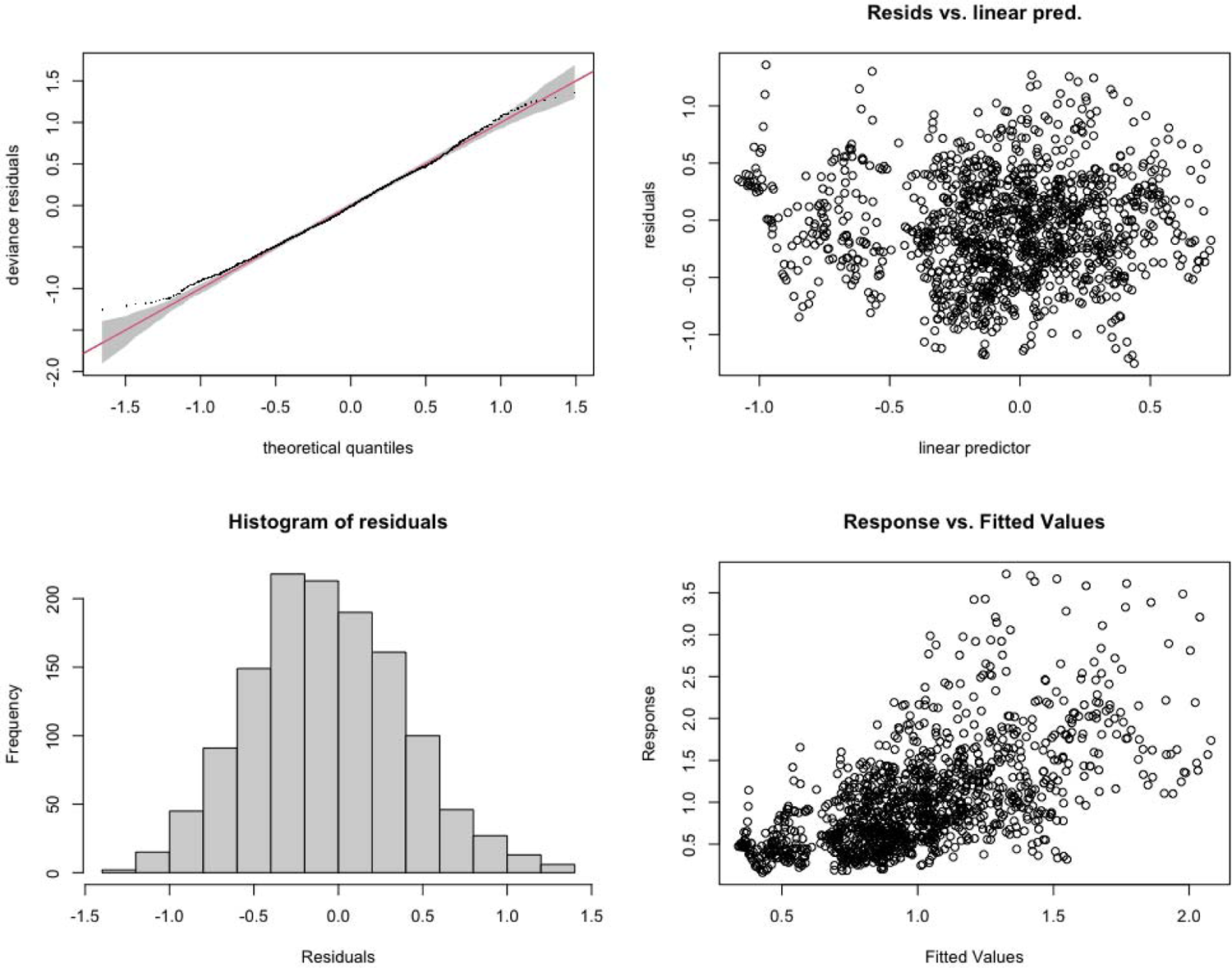
Model diagnostic plots for Model Delta-RCIR. The correlation coefficient between fitted RCHR and predicted RCHR is 0.61.

**Appendix figure S13:**
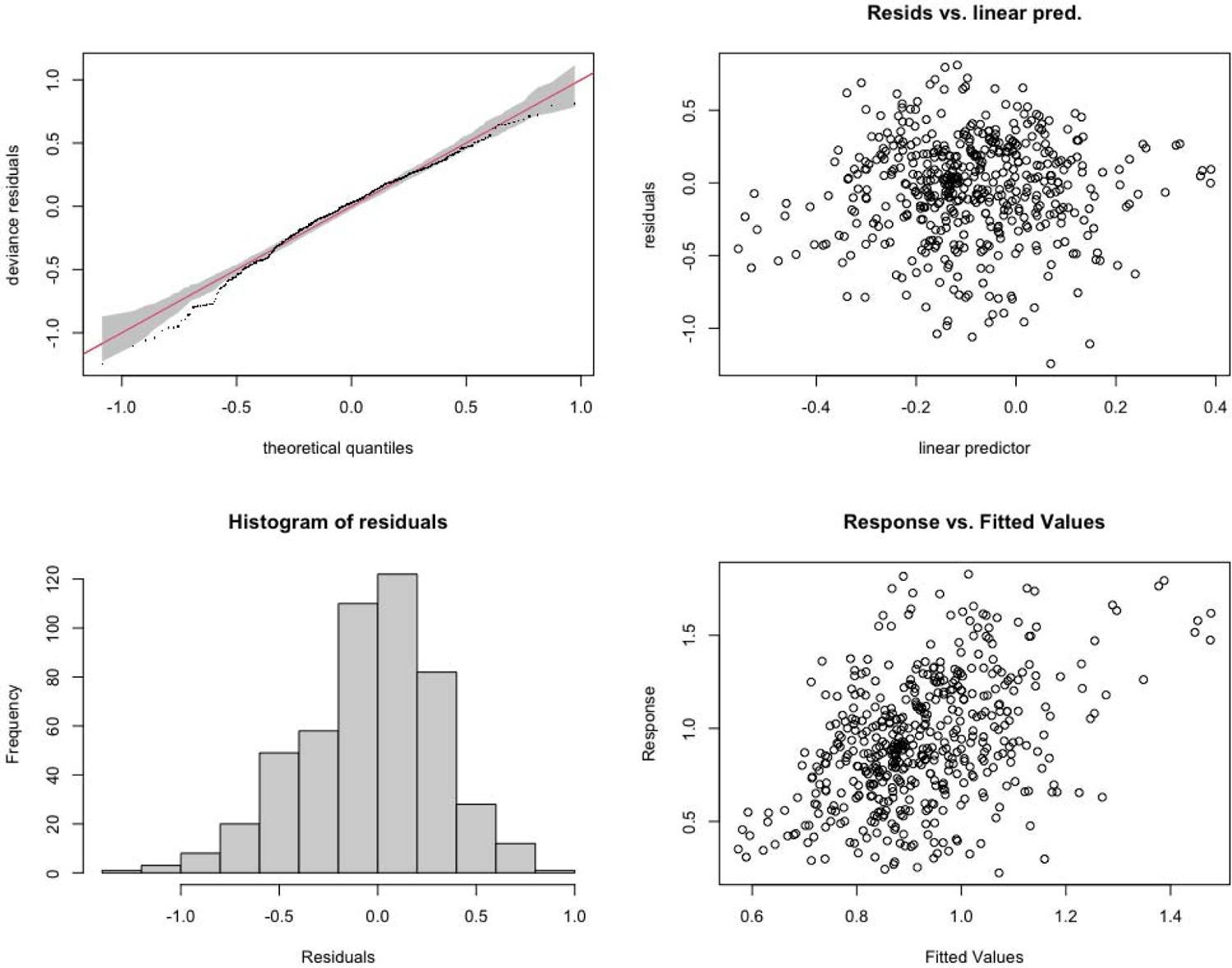
Model diagnostic plots for Model Omicron-RCIR. The correlation coefficient between fitted RCHR and predicted RCHR is 0.43.

**Appendix figure S14:**
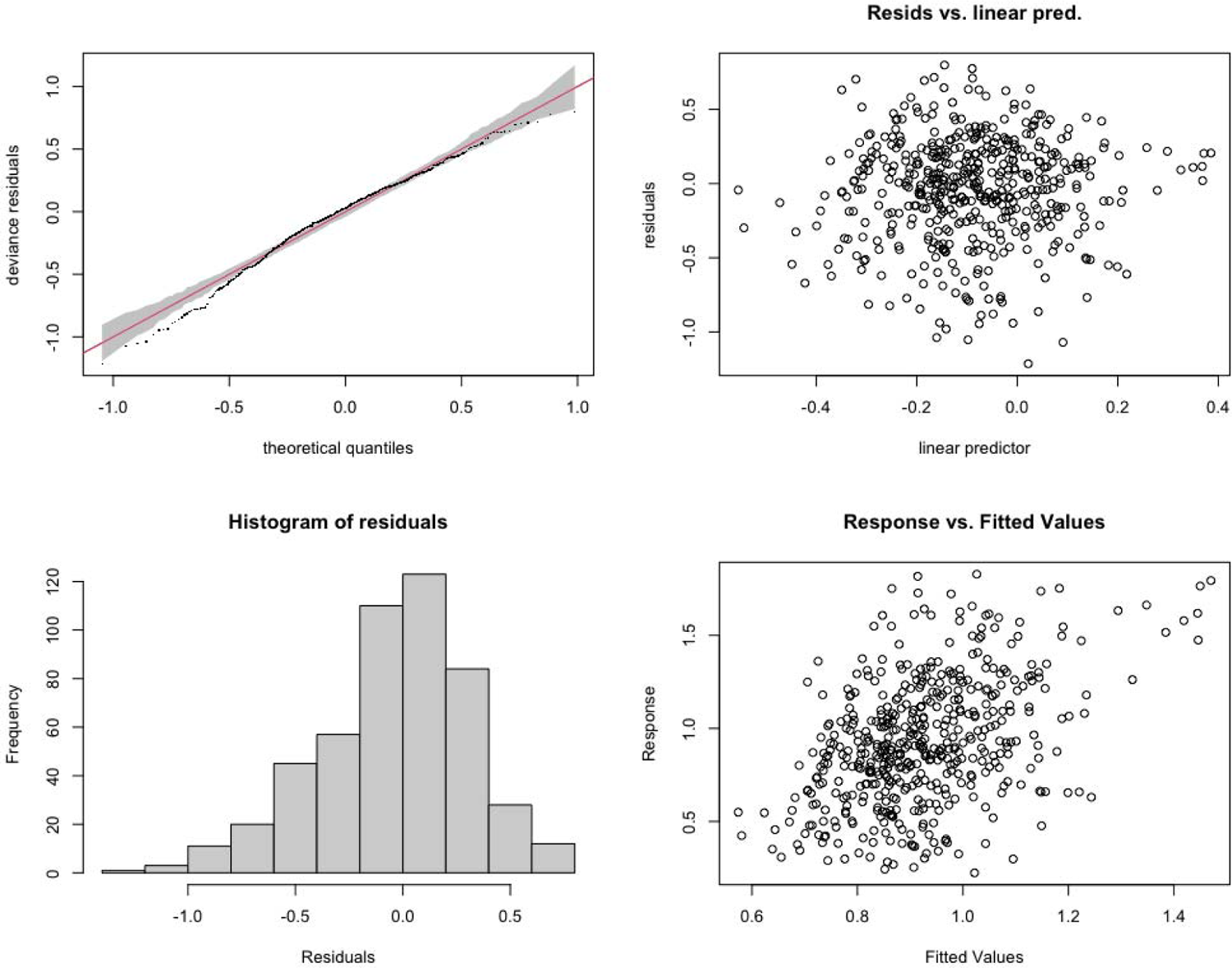
Model diagnostic plots for Model Omicron-Booster-RCIR. The correlation coefficient between fitted RCHR and predicted RCHR is 0.44.

